# Amplicon-based targeted next generation sequencing (tNGS) for *Mycobacterium tuberculosis* - unpacking the ‘black box’

**DOI:** 10.64898/2026.09.27.26364088

**Authors:** Xiaomei Zhang, Eby M. Sim, Connie Lam, Andrea Bustamante, Taryn Crighton, Ben J Marais, Vitali Sintchenko

## Abstract

The WHO recently endorsed two commercial culture-free targeted next-generation sequencing (tNGS) assays for detecting drug resistant Mycobacterium tuberculosis. These proprietary tNGS platforms present a ‘black box’. We developed an in-house tNGS assay using Oxford Nanopore Technology sequencing to detect an expanded range of resistance mutations directly from clinical specimens. The assay targeted 44 genomic loci; hsp65 for Mycobacterium tuberculosis complex (MTBC) identification; 12 loci for MTBC lineage classification; and 31 drug resistance loci predicted resistance to 16 anti-TB drugs. In total, 98 amplicons were distributed across three PCR pools. Analytical sensitivity was assessed using M. tuberculosis H37Rv, and proof-of-concept performance was evaluated in 63 archived clinical specimen extracts and one ‘real-life’ sputum specimen in comparison with Illumina whole genome sequencing (WGS). All amplicons were successfully amplified and sequenced from M. tuberculosis H37Rv genomic DNA. At ≥100 genome copies/μL, 71.4% of amplicons met ≥20( mean depth across six replicates, including all loci for rifampicin, ethambutol, pyrazinamide, fluoroquinolone and bedaquiline resistance. In 63 clinical specimens, (90% of amplicons achieved a mean depth of (20( in specimens with IS6110 PCR cycle threshold (Ct) values ≤24. With Ct value ≤27 lineage classification was concordant with WGS in 94.7% (18/19) of specimens and drug-susceptibility prediction showed 94.7% (18/19) agreement with both phenotypic and genotypic (WGS) testing. In a single clinical case (Ct 26.5), lineage and resistance-associated mutations were accurately identified directly from sputum. The findings provide new insights into rapid culture-independent tNGS approaches for rapid M. tuberculosis drug resistance detection with lineage assignment.

## Introduction

The timely and accurate identification of mutations associated with drug resistance in *Mycobacterium tuberculosis* is critical for guiding effective treatment regimens and limiting the ongoing transmission of drug-resistant tuberculosis [1, 2]. The second edition of the WHO Catalogue identified ∼1,370 mutations (Groups 1 and 2) associated with resistance to 15 antituberculosis drugs [2]. Nucleic acid amplification tests (NAAT), such as the GeneXpert MTB/RIF and line probe assays, are well established and endorsed by the World Health Organization (WHO) as methods to detect common resistance-conferring mutations [3]. However, these assays are limited in both the number and diversity of mutations they detect, targeting only a small subset of drugs and resistance loci included in the WHO Catalogue.

In July 2023, the WHO issued guidance supporting the use of two commercial rapid targeted next-generation sequencing (tNGS) assays for the diagnosis and drug resistance profiling of *M. tuberculosis*, directly from clinical specimens [4]. tNGS offers several advantages over NAAT, including potential improved sensitivity for low-frequency variants and the ability to detect mixed infections with discordant resistance profiles [5]. Importantly, tNGS panels can be designed to interrogate a broader range of resistance determining regions and mutation types, including insertions, deletions, frameshifts, and variations in repetitive regions that contribute to clinically relevant resistance. A recent meta-analysis surmised that tNGS approaches have high sensitivity (>95%) for isoniazid (INH), rifampicin (RIF), and ethambutol (EMB) and high specificity (>95%) for most other antituberculosis drugs, except ethambutol [6].

Another advantage of tNGS is its ability to bypass the need for culture, enabling genomic analysis directly from clinical specimens. Culture independent tNGS approaches offer a faster alternative to whole genome sequencing (WGS), which requires pre-culture for reliable performance [7]. Several commercial tNGS panels have shown potential in delivering drug resistance profiles in patients with high bacillary loads [8]. However, performance with lower bacillary loads are not well characterised, and the proprietary nature of commercial tNGS panels can create a ‘black box’ where end-users are unable to interrogate the panel and adapt the technology. This can impede critical assessment and adaptation of panels to regional-specific resistance patterns or to newly described drug resistance mutations.

In response, we aimed to develop and test a flexible in-house tNGS assay with a fully transparent and adaptable assay design for rapid detection of resistance-associated mutations and lineage markers in *M. tuberculosis* directly from clinical specimens using Oxford Nanopore Technology (ONT) long read sequencing. Target selection was guided by the second edition of the ‘WHO Catalogue of Mutations in *Mycobacterium tuberculosis* Complex and Their Association with Drug Resistance’[2], including loci associated with resistance to drugs included in the new BPaL (bedaquiline, pretomanid, linezolid) regimen [9]. We assessed the performance of this assay by determining its limit of detection and its ability to be used on clinical specimens with different bacterial loads. We also evaluated concordance with culture-based phenotypic drug susceptibility testing (pDST) and WGS.

## Methods

### Amplicon design and multiplex PCR pools

This study targeted 44 genomic loci (Table S1), including (i) 31 loci associated with drug resistance to 16 anti-tuberculosis drugs listed in the 2023 WHO Catalogue [2], (ii) *hsp65* for *M. tuberculosis* complex (MTBC) identification [10, 11], and (iii) 12 genomic loci related to phylogenetic lineage classification (Lineages 1 to 9) and differentiation for some members within the MTBC, including *M. bovis* (La1), *M. caprae* (La2), and *M. orygis* (La3) [12]. The panel was designed to include resistance-associated regions covering WHO Catalogue group 1 and group 2 mutations, as well as the majority of group 3 mutations available at the time of assay design, while retaining the flexibility to update or expand targets as new resistance-associated mutations are identified.

Primers were designed using PrimalScheme v3.0.2 (https://github.com/artic-network/primalscheme3) with the High-GC model and a minimum base frequency threshold of 0.01. Amplicons were approximately 450 bp in length to ensure consistent and efficient amplification. Due to both gene length and location of mutational hotspots, the 31 drug resistance-associated loci were covered by multiple overlapping primer pairs, resulting in a total of 85 primer pairs. With species and lineage-related regions included, our tNGS panel included a total of 98 primer pairs (Table S2).

Primer pairs were evaluated using MFE-primer [13] with default setting to assess potential primer-dimer formation, cross-reactivity, non-specific amplification, and secondary structures against *M. tuberculosis* H37Rv reference genome (NCBI RefSeq: NC_000962.3). Primer pairs were grouped based on predicted compatibility, with manual adjustments to minimize dimer formation and prevent cross-reactivity (Table S2). For targets with multiple amplicons, overlapping primer pairs were assigned to separate PCR pools to minimise primer-dimer formation, reduce amplification interference between overlapping amplicons, and improve amplification efficiency across the highly multiplexed assay. Based on these assessments, the 98 primer pairs were distributed into three non-overlapping pools (Pool, 1 n=34; Pool 2, n=32; Pool 3, n=32). All primers were synthesized with standard desalting purification (Sigma-Aldrich, Oceania Pty Ltd).

### Genomic DNA extraction

DNA from the *M. tuberculosis* H37Rv reference strain was extracted as previously described [14] and quantified (Qubit dsDNA HS Assay, Thermo Fisher Scientific). Clinical specimens underwent routine decontamination prior to nucleic acid extraction as part of standard mycobacterial diagnostic processing. DNA from clinical specimens were extracted using the NucliSENSE EasyMag total nucleic acid extraction system (BioMerieux) after all routine diagnostic testing was performed; residual extracts were stored at -20°C.

### Multiplex PCR reaction, library preparation and sequencing

DNA from *M. tuberculosis* H37Rv was used for calibration. Positive and negative controls were included throughout assay optimisation and sequencing experiments. PCR reactions of 25 μL were set up using the Q5^®^ High-Fidelity DNA polymerase and its associated reagents (New England Biolabs), including addition of the 5X Q5 High GC Enhancer, according to the manufacturer’s instructions. Final primer concentrations for each of the three pools are provided in Table S2. PCR cycling conditions were as follows: initial denaturation at 98°C for 5 min; 40 cycles of 98°C for 30 s, 66°C for 30 s, 72°C for 60 s; final extension at 72°C for 5 min.

PCR products from the three pools were combined into a single pool and purified using AMPure XP beads (Beckman Coulter) at a 1:1 bead-to-sample volume ratio. Purified amplicons were quantified using the Qubit dsDNA High Sensitivity Assay Kit (Invitrogen) and normalized to 120 ng per sample prior to library preparation. Libraries were prepared using the Oxford Nanopore Technologies (ONT) Rapid Barcoding Kit 96 V14 (SQK-RBK114.96) loaded onto an R10.4.1 flow cell and sequenced on a MinION Mk 1b. Real-time base calling and demultiplexing on the HAC model was performed using MinKNOW v24.11.10. Run times were variable and dependent upon data acquisition, with sequencing run terminated when all samples would have acquired enough data to cover the tNGS panel to a theoretical depth of 100×, when monitored on the MinKNOW app.

### Analytical sensitivity analysis using H37Rv

The limit of detection was assessed using 10-fold serial dilutions of *M. tuberculosis* H37Rv DNA, quantified using the Qubit dsDNA High Sensitivity Assay Kit and subsequently diluted to 250 pg/μL, 25 pg/μL, 2.5 pg/μL, 250 fg/μL, and 25 fg/μL. For each dilution, 5 μL was added to each PCR pool as described above, corresponding to approximately 10,000, 1000, 100, 10, and 1 genome copies/μL in the final reaction mixture. Each dilution was tested in triplicate with two independent runs to evaluate reproducibility of amplification and sequencing performance across decreasing genome copy concentrations.

Analytical sensitivity was evaluated based on amplification success and sequencing consistency with mean amplicon sequencing depth across all targeted regions. Successful target detection was defined as achieving a mean amplicon sequencing depth ≥20× across the targeted amplicons and achieving ≥90% concordance in amplicon reads. This predefined assay performance threshold was distinct from the minimum 10× read depth threshold used by TB-Profiler v6.6.4 [15] for mutation detection and variant calling. The corresponding genome copy number per μL was calculated using the formula [16] [(DNA mass in grams × 6.02 × 10^23^) / (Genome size in bp × 660)]; using the *M. tuberculosis* H37Rv genome size (4.4 Mb).

### Proof-of-concept evaluation using clinical specimens

To assess ‘real life’ diagnostic potential, a proof-of-concept evaluation was performed on 63 clinical specimens at the Institute of Clinical Pathology and Medical Research (ICPMR) NSW Health Pathology, collected between 1 April 2022, and 31 March 2024. As this initial proof-of-concept study was designed to evaluate key operating parameters and assess assay performance characteristics, the sample size was determined by the availability of clinical specimens with both pDST and WGS-based genotypic DST (gDST) results during the study period. All 63 samples were split across two flow cells with 31 and 32 samples respectively. Both flow cells were run for 23 hours. Of these, 47 (74.6%) were respiratory specimens, including 38 sputum specimens (60.3%) and nine bronchoalveolar lavages (14.3%) [17]. Complementary data included real-time PCR cycle threshold (Ct) values targeting IS*6110* obtained from routine clinical diagnostic testing records, pDST, and WGS; if successfully performed (Figure S1).

Concordance was used to assess agreement between the novel tNGS assay and reference methods (pDST and/or WGS-based gDST) for evaluation of drug susceptibility, and was calculated as n/N, where n is the number of specimens with matching results and N is the total number of specimens with available reference results.

### Bioinformatic analysis

Sequencing reads were separately aligned to two references using Minimap2 [18]: i) a custom reference consisting of the concatenated sequences of all 98 amplicons (Supplementary Data), and ii) the *M. tuberculosis* H37Rv reference genome (NCBI RefSeq: NC_000962.3) to evaluate drug resistance prediction using the 2023 WHO catalogue [2]. Parsing of alignments and alignment statistics, which included number of mapped reads, mean depth, and coverage metrics, were performed using SAMtools v1.17 [19]. All resulting BAM files were visualised using Integrative Genomics Viewer (IGV)[20] to assess alignment quality. Sequencing depth across amplicons and resistance-associated nucleobase (henceforth referred to as genomic markers) included in the WHO Catalogue was evaluated for all clinical specimens.

To call a mutation associated with either drug resistance or lineage assignment, the implicated amplicon and genomic markers required a mean read depths of ≥ 20× and ≥10×, respectively. Lineage and drug resistance were classified using TB-Profiler v6.6.4 [15]. Minority variants, defined as having an allelic frequency (AF) of 3-49% supported by ≥4 reads [21], were detected using LoFreq v2.1.1 [22].

## Results

### tNGS assay performance - using M. tuberculosis H37Rv

With DNA extraction from *M. tuberculosis* H37Rv (DNA concentration: 0.25 ng/μL), all 98 targets were successfully amplified and sequenced; 80,470 mapped reads with average of 821 reads per amplicon (SD 418; median 776, IQR 505 – 1097). The mean read depth across all amplicons was 488× (SD 261; median 461, IQR 283 – 652) and 98.0% of amplicons achieved a mean depth of ≥20×; 2 amplicons had mean depths of 16× and 19×, respectively (Figure S2). The median mapping quality score (mean MAPQ) was 56.4 (IQR 55.0 – 56.9), indicating high-confidence alignment. All 1,278 markers listed “Associated with Resistance” (Group 1) or “ Associated with Resistance – Interim” (Group 2) in the 2023 WHO Catalogue [2] were captured in our tNGS panel. Sequencing depth across 14 markers for species and lineage identification ranged from 31× to 1552×, with an overall average of 523×.

Figure 2 shows the analytical sensitivity of the tNGS assay with diminishing DNA input, evaluated using six replicates of *M. tuberculosis* H37Rv. At inputs of ≥100 genome copies/μL, the average sequencing mean depth was >100× for amplicons and genomic markers, declining to 55× for amplicons and 62× for genomic markers with 10 genome copies/μL (Figure 2a). With a DNA input of ≥100 genome copies/μL, >95% of genomic markers met the ≥10× depth threshold, declining to ≥82% with 10 genome copies/μL.

**Figure 1:**
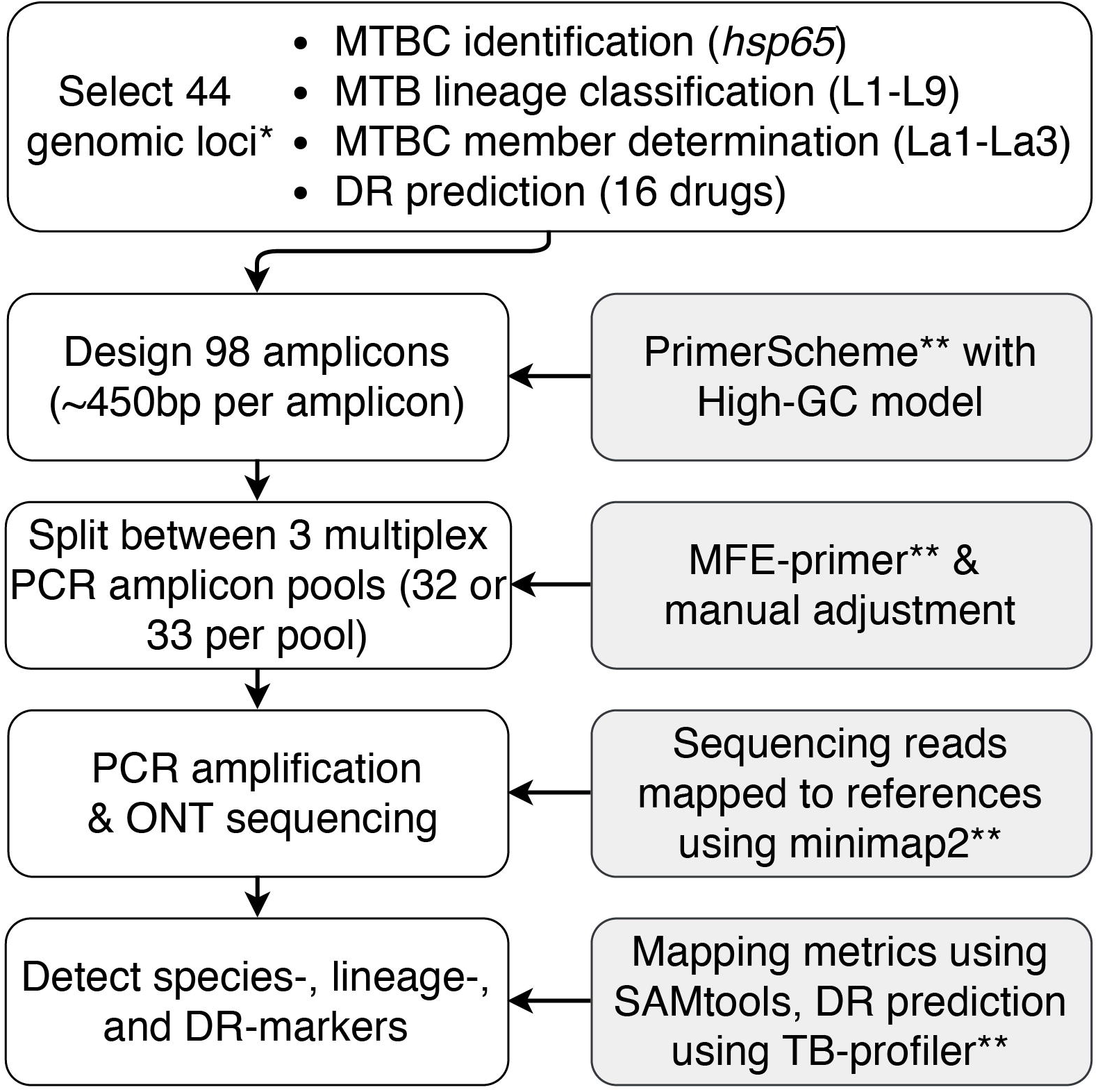
Flowchart summarising tNGS assay development. DR: drug resistance; L: lineage; La: animal-adapted lineage of the MTBC; MTBC: *Mycobacterium tuberculosis* complex; PCR: Polymerase Chain Reaction; ONT: Oxford Nanopore Technology; TB: tuberculosis; tNGS: Amplification-based targeted next generation sequencing. *Genomic loci are listed in Table S1; **Detailed methods are described in the Materials and methods section.

**Figure 2:**
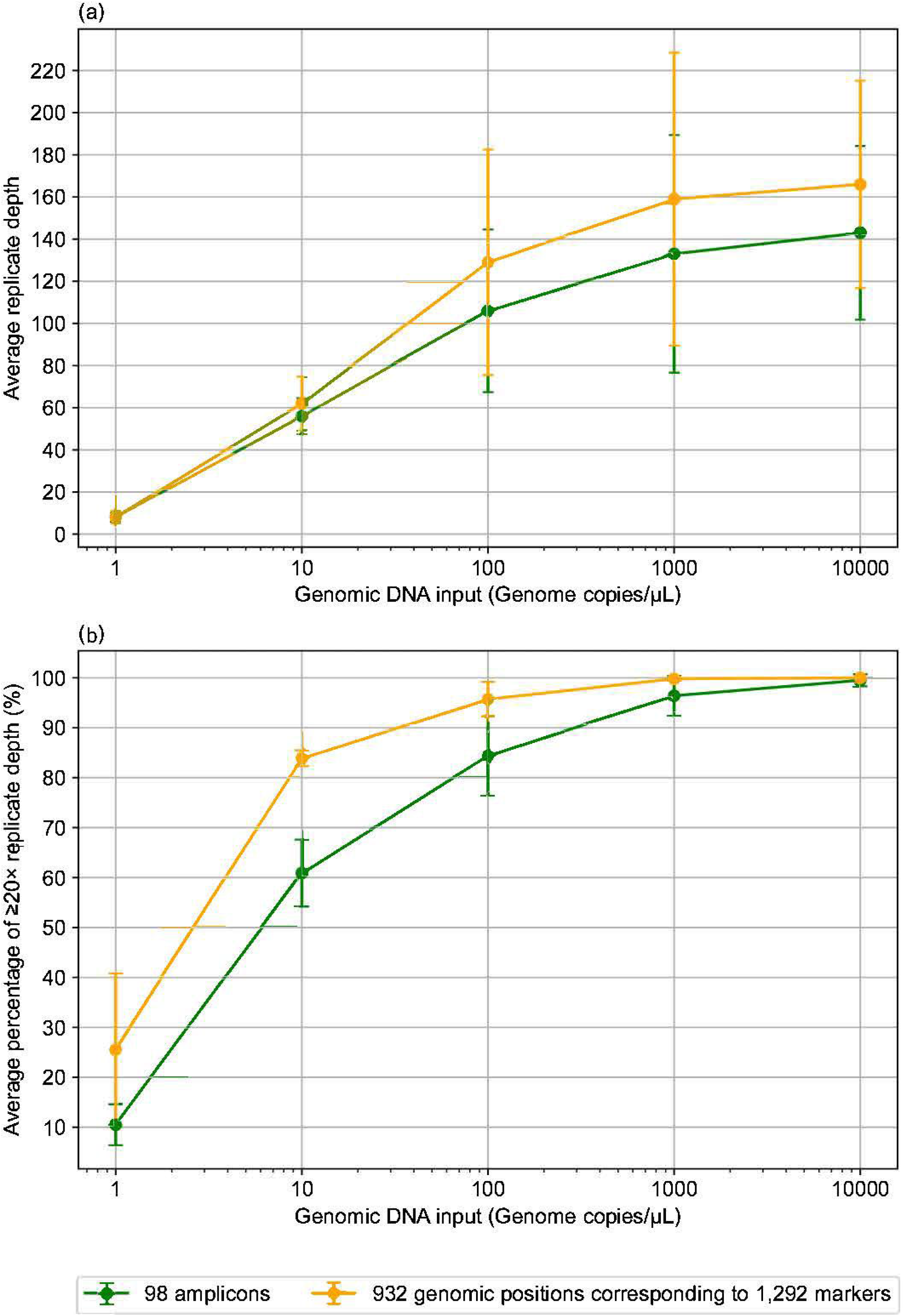
tNGS assay detection sensitivity with increasing amounts of *M. tuberculosis* H37Rv genomic DNA. (a) Average amplicon (mean depth; green) and genomic marker depth (orange). (b) Average percentage of amplicons and genomic markers reaching ≥20× mean depth across six replicates. Bars indicate the standard deviation (SD). tNGS: Amplification-based targeted next generation sequencing.

tNGS assay sensitivity to detect specific drug resistance markers was dependent on *M. tuberculosis* genome copy number (Figure 3). At ≥100 genome copies/μL, 91.5% of genomic markers met the depth threshold (Figure 3a). With an input of ≥100 genome copies/μL most genomic markers associated with resistance to rifampicin, ethambutol, pyrazinamide, fluoroquinolones (levofloxacin/moxifloxacin), and bedaquiline were detected at ≥10× depth (Figure 3b). We estimate that ∼100 genome copies/μL (∼0.5 pg *M. tuberculosis* DNA) are required to reliably amplify >95% of the targeted regions. Figure S3 shows over 71.4% amplicons consistently achieved ≥20× mean depth in all six replicates at ≥100 genome copies/μL. Amplicons and genomic markers that did not meet the ≥20× mean depth threshold at DNA input levels of 100, 1,000, and 10,000 genome copies/μL are listed in Table S3.

**Figure 3:**
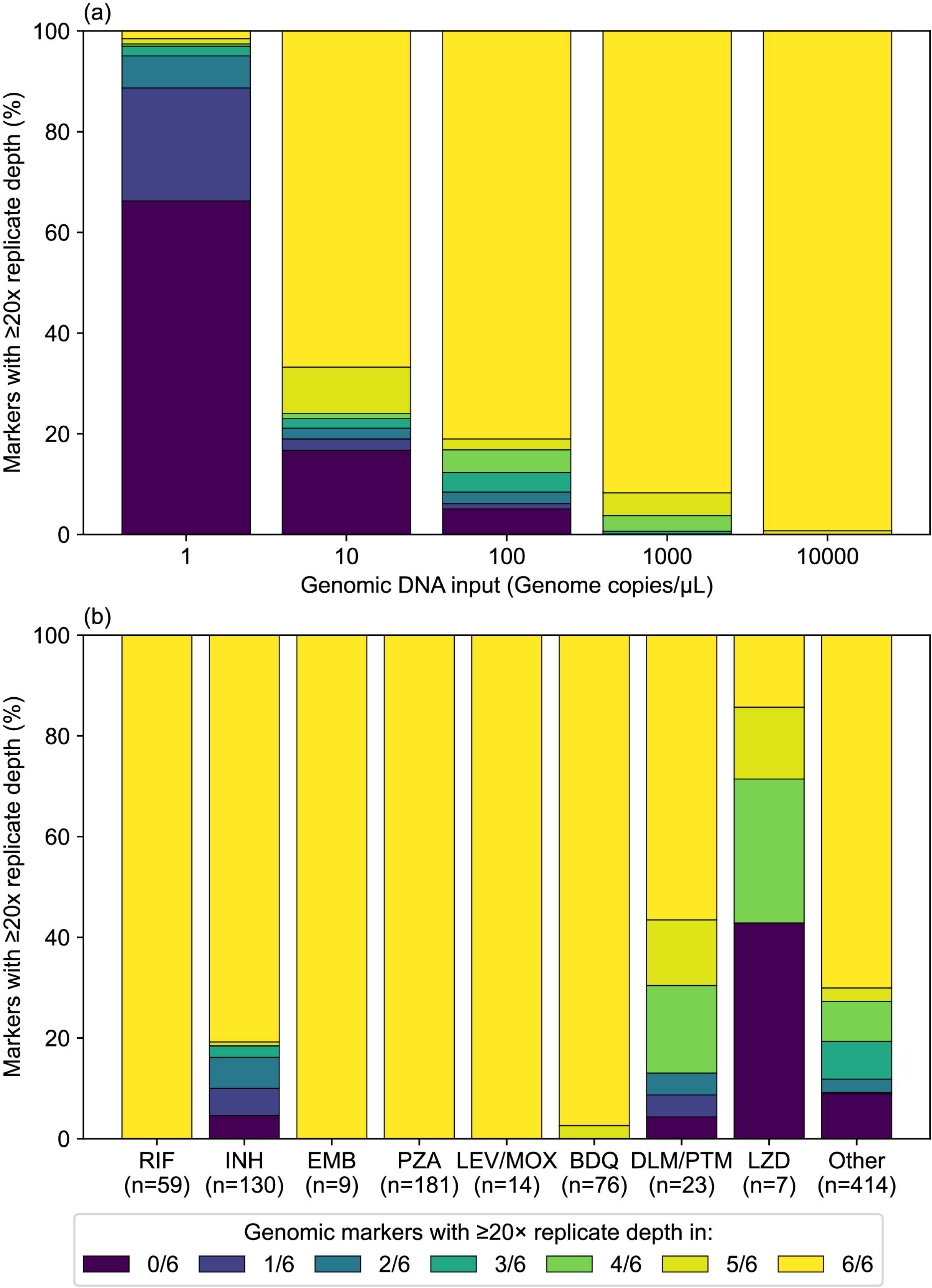
tNGS assay detection sensitivity with increasing amounts of *M. tuberculosis* H37Rv genomic DNA, focused on genomic markers associated with resistance to key drugs. (a) Detection of 932 genomic positions corresponding to 1,292 target genomic markers, including species and lineage (14 markers) and drug resistance (DR) (1,278 markers). (b) Focus on 918 genomic positions (1,278 DR markers) consistently detected (≥10 depth across six replicates) with DNA input of ≥100 genome copies/μL. *Total number of genomic positions evaluated per drug displayed on the x-axis in brackets. BDQ: bedaquiline; DLM: delamanid; EMB: ethambutol; INH: isoniazid; LEV: levofloxacin; LZD: linezolid; MOX: moxifloxacin; PTM: pretomanid; PZA: pyrazinamide; RIF: rifampicin; Other: ethionamide, amikacin, capreomycin, kanamycin, streptomycin; tNGS: Amplification-based targeted next generation sequencing.

### tNGS assay performance - using clinical specimens

Among 63 clinical specimens, those with PCR Ct values ≤24 (n=4) achieved mean amplicon coverage of ≥99.9%, with at least 91.8% of the amplicons reaching a mean depth of ≥20× (Figure S4). For specimens with Ct values between 24 and 27 (n=15), the mean amplicon coverage was ≥97.7%, and 85.3% of amplicons met the ≥20× mean depth threshold. In specimens with Ct values between 27.1 and 33 (n=19), the mean depth was inconsistent across amplicons but some drug resistance inferences could still be made, which became impossible from specimens with Ct values >33 (n=25).

Figure 4 shows sequencing depth for Group 1 (“Assoc w R” in the 2023 WHO Catalogue) genomic markers associated with drugs of highest clinical relevance. All specimens with Ct values ≤24 demonstrated sufficient sequencing depth for drug resistance prediction (e.g., TB-Profiler depth threshold 10×), 93.3% (14/15) of specimens with Ct values between 24 and 27 and only 36.8% (7/19) with Ct values >27. Mean amplicon sequencing depth and sequencing depth across resistance-associated markers included in WHO Catalogue groups 1 and 2 for first- and second-line drugs is provided in Table S4 and Table S5, respectively. Table 1 shows drug resistance detection concordance of between the novel tNGS assay and reference phenotypic and WGS-based genotypic drug susceptibility testing (p and gDST) for drug resistance detection. Based on pDST and/or gDST, 9/63 (14.3%) specimens were drug resistant; 2 multidrug (MDR; resistance to isoniazid and rifampicin) and 7 mono-drug resistant; 4 isoniazid and one each resistant to ethambutol, streptomycin and a fluoroquinolone. Among specimens with Ct ≤27 (n=19), tNGS had 94.7% (18/19) concordance with both pDST and gDST. No minority variants associated with drug resistance were detected in any clinical specimens.

**Figure 4:**
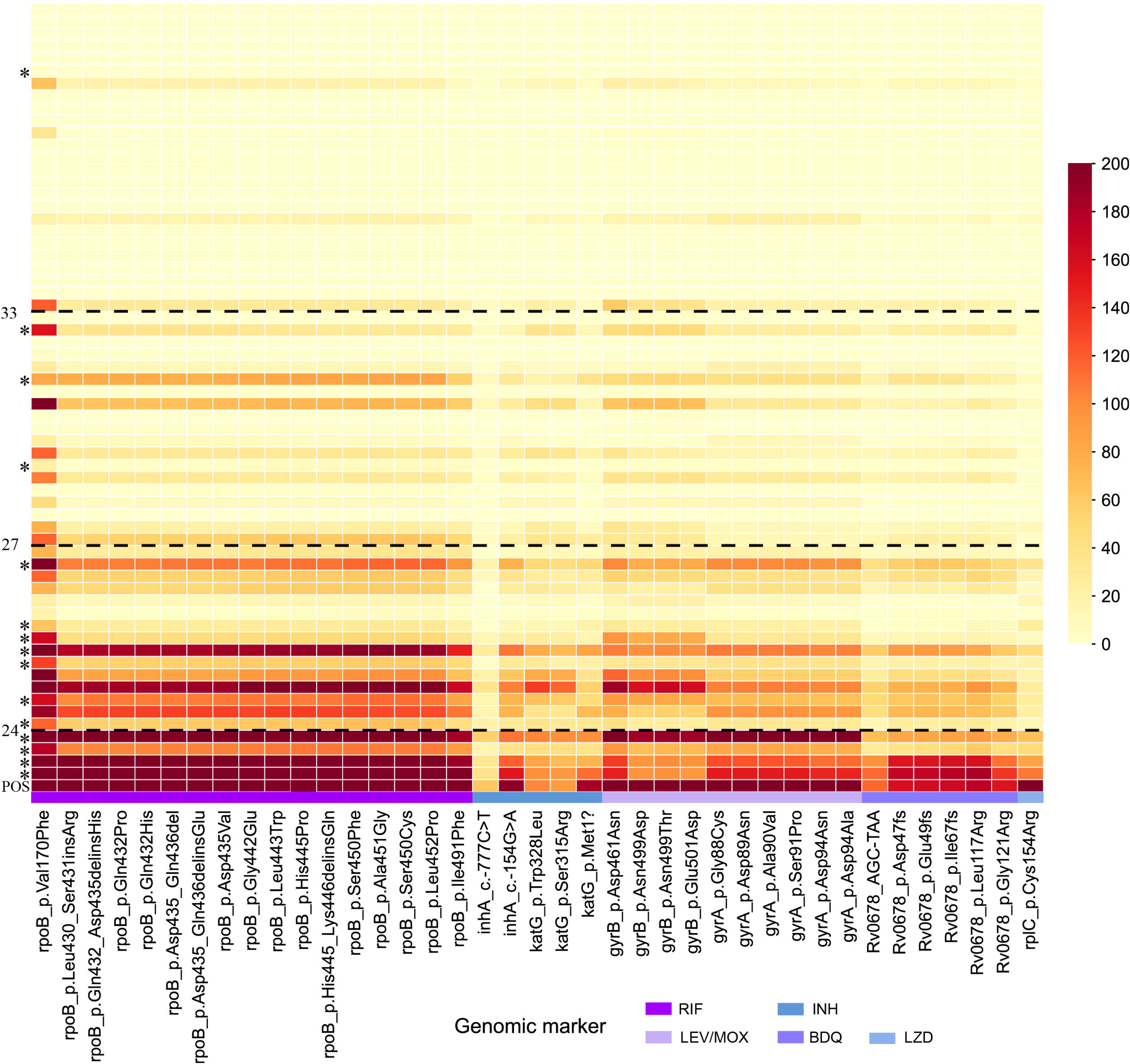
tNGS assay detection sensitivity in 63 clinical specimens, focused on key first-line and BPaL/M drugs. Each column represents a drug-resistant marker classified as “Assoc w R” (group 1) in the 2023 WHO Catalogue**. Each row represents a clinical specimen (n=63), ordered by increasing IS*6110* PCR Ct value (decreasing DNA amount). A positive control (POS) of 10,000 genome copies/μL is included - first row. *Smear-positive specimens. **No Group 1 genomic markers described of pretomanid resistance. Dashed line: IS*6110* PCR Ct value thresholds according to amplicon mean depth ≥20×: • High clinical utility – Ct value ≤24; ≥90% (average 91.8%) • Some clinical utility – Ct value 24 - 27: 50-90% (average 63.3%) • Limited clinical utility – Ct value 27.1 - 33: <50% (average 22.8%) • No clinical utility – Ct value >33 BPaL/M: bedaquiline, pretomanid, linezolid, moxifloxacin; BDQ: bedaquiline; Ct: real-time PCR cycle threshold values targeting IS*6110*; INH: isoniazid; IS: Insertion Sequence; LEV: levofloxacin; LZD: linezolid; MOX: moxifloxacin; PCR: Polymerase Chain Reaction; RIF: rifampicin; tNGS: Amplification-based targeted next generation sequencing; WHO: World Health Organization.

**Table 1:** Drug resistance detection concordance of tNGS assay with pDST and WGS-based gDST, relative to the IS*6110* PCR Ct value.

| IS6110 PCR Ct value (N) | Drug susceptible *M. tuberculosis* (DS) |  | Drug resistant *M. tuberculosis* (DR) |  | Combined concordance with pDST and/or gDST, % (n/N) |
| --- | --- | --- | --- | --- | --- |
|  | DS by pDST and/or gDST, N | Concordance with pDST and/or gDST, % (n/N) | DR by pDST and/or gDST, N | Concordance with pDST and/or gDST, (n/N) |  |
| ≤27 (19) | 16 | 100 (16/16) | 3 | 2/3* | 94.7 (18/19)* |
| 27.1 - 33 (19) | 17 | 35.3 (6/17)** | 2 | 1/2** | 36.8 (7/19)** |
| > 33 (25) | 21 | 14.3 (3/21)*** | 4 | 1/4*** | 16.0 (4/25)*** |
| Total (63) | 54 | 46.3 (25/54) | 9 | 4/9 | 46.0 (29/63) |

Percentages are not reported for categories with small sample sizes (N < 10; results are presented as counts (n/N) only.
\* Insufficient coverage and sequencing mean depth in one specimen - 3,075 mapped reads with only 23.5% (23/98) of amplicons reaching ≥20x mean depth (range: 0-121x).
\*\* Insufficient coverage and sequencing mean depth in 12 specimens.
\*\*\*Insufficient coverage and sequencing mean depth in 21 specimens.
Ct: real-time PCR cycle threshold values targeting IS6110; gDST: genotypic drug susceptibility testing; INH: isoniazid; IS: Insertion Sequence; PCR: Polymerase Chain Reaction; pDST: phenotypic drug susceptibility testing; RIF: rifampicin; tNGS: Amplification-based targeted next generation sequencing; WGS: Whole genome sequencing.

The sequencing depth for the broader set of 130 markers conferring resistance to ethambutol, pyrazinamide, ethionamide, amikacin, capreomycin, kanamycin, streptomycin, as well as 14 lineage-markers is presented in Figures S5 and S6. All specimens with PCR Ct values of ≤27 (n=19) had near universal species and lineage coverage (Table S6). *M. tuberculosis* lineage classification (lineage 1-4) was concordant with WGS results in 18/19 (94.7%) specimens; one specimen (Ct 26.8) had low performance with only 35.7% (35/98) of amplicons reaching ≥20× mean depth. No hetero-resistance or mixed populations were detected in any of the clinical specimens tested, consistent with culture-based pDST results (data not shown). From these non-host depleted samples the proportion of MTBC reads, as assessed by Centrifuge v1.0.4.2 [23] against the p_compressed+h+v database, ranged from >0% to 58.4% of the total data (Table S7). The mapper used within TB-Profiler for ONT reads only works well between sequences with divergence of ∼15.0%. While spurious mapping could be possible, the general sequence divergence between *M. tuberculosis* and human sequence would make this highly unlikely. In the run configuration used for the 63 samples, the cost per sample would be approximately AUD $ 58 (Figure S7)

### tNGS utility for expanded drug resistance screening

The tNGS panel was also assessed for its value add in complementing current testing algorithms. DNA was extracted from a sputum specimen, not assessed in the 63 specimen set, provided by a patient with drug resistant TB detected by both Xpert-MTB/RIF and Xpert-MTB/XDR; IS*6110* Ct value 26.5. An entire R10.1 flow cell was dedicated to this run. Within six hours a *M. tuberculosis* lineage 4 strain was identified with predicted resistance to rifampicin, isoniazid, ethambutol, pyrazinamide, moxifloxacin/levofloxacin and ethionamide (TB-profiler). All amplicons had ≥35× mean depth; average 196×. Mutations covered by both Xpert MTB/RIF and MTB/XDR included *rpoB*_p.Ser450Leu (AF: 0.96; read depth: 196×), *ahpC*_c.-81C>T (AF: 0.96; read depth; 67×) and *katG_*p.Ser315Thr (AF: 0.99; read depth: 129×). For first line drugs not covered by Xpert, the Group 1 mutations for ethambutol and pyrazinamide resistance were *embB*_p.Met306Val (AF: 0.98; read depth: 298×) and *pncA*_p.Val139Ala (AF: 0.98; read depth: 248×) respectively. For second line drugs, the associated Group 1 mutations were *gyrA*_p. Gly88Cys (AF: 1; read depth: 223×) and *ethA*_c.341dupA (AF: 1; read depth 169×) for moxifloxacin/levofloxacin and ethionamide respectively. When the associated culture was sequenced, WGS results corroborated findings from tNGS panel. When used in this run configuration, the cost per sample was approximately AUD $1619 (Figure S7).

## Discussion

Our results demonstrate excellent tNGS assay performance in specimens with very high bacterial loads (Ct ≤24) and adequate performance in those with high bacterial load (Ct ≤27). Our estimate that a minimum of ∼100 genome copies/μL are required to reliably amplify targeted regions is comparable to a reported limit of detection of ∼100 genome copies (0.45 pg) for probe based *M. tuberculosis* target enrichment panel [24], but is much less sensitive than the ∼4.5 genome copies required by Xpert-MTB/RIF [25]. IS*6110* real-time PCR Ct values were used as a practical indicator of relative *M. tuberculosis* DNA abundance within the study cohort. However, variation in IS*6110* copy number across *M. tuberculosis* lineages may influence the relationship between Ct value and bacterial load in different epidemiological settings.

Sputum specimens with lower *M. tuberculosis* DNA content (Ct values >27) yielded incomplete or undetermined results, including reduced MTBC species identification performance. However, Ct value did not always predict tNGS amplification performance, as some specimens with relatively high Ct values still achieved adequate amplification and sequencing depth. This variability likely reflects multiple factors beyond relative *M. tuberculosis* DNA abundance alone, including specimen quality, DNA integrity, repeated freeze-thaw cycles, and variation in IS*6110* copy number across strain backgrounds. Some variability in amplicon sequencing depth was also observed across primer pairs, which is expected in highly multiplexed PCR assays targeting GC-rich *M. tuberculosis* genomic regions despite extensive *in silico* optimisation and multiplex pool adjustment. However, the majority of targeted regions exceeded the predefined 20× mean sequencing depth threshold for successful target detection. High Ct value specimens may be partially attributed to the re-use of archived specimens employed in this study, which had undergone multiple rounds of routine testing for diagnostic purposes with repeated freeze-thaw cycles potentially compromising specimen integrity. However, the high mean sequencing depth (average of 171× across all amplicons) achieved in a ‘real life’ TB case (Ct value 26.5) suggests that analytical sensitivity may be higher if fresh clinical specimens are used. Adding additional enrichment steps or improved human DNA depletion may further enhance test sensitivity.

We detected drug resistance associated mutations in 31 loci specified by the WHO Catalogue, but no new variants within known genes of interest. This reflects the relatively small number of clinical specimens tested and the low rates of drug resistance in the study setting. Novel genomic regions could not be interrogated, which is an inherent limitation of all tNGS assays. However, the in-house assay design allows individual targets to be examined and enables target regions to be modified or expanded as additional clinically relevant resistance-associated mutations are identified and verified.

Mixed bacterial populations have been recognised as potential drivers of treatment failure [26, 27] and tNGS may detect minority variants directly from clinical specimens, which is attractive since it avoids the strain selection that occurs during culture. The commercial Deeplex-MTB tNGS assay can detect heteroresistance at a limit of >6% minority populations [28]. We previously demonstrated the presence of mixed and heteroresistant populations in cultured specimens using ≥4 Illumina WGS reads for minority variant detection [21], which suggests that the detection of ∼5% minority variants should be achievable with our in-house tNGS assay. However, no minority variants were detected in the current study. Controlled mixing studies using defined mixed bacterial populations were also not performed and required to further define the limit of detection for heteroresistance and mixed infections. The detection of minority variants <5% should be interpreted with caution considering the intrinsic error rate of ONT R10.4.1 sequencing.

Drug resistance determination directly from clinical specimen reduces the cost, time and resources associated with culture-based testing. Thus, tNGS may provide benefits in regions which have limited or unreliable access to culture-based testing. A recent cost-effectiveness study of tNGS use for TB drug resistance in low and middle income countries provided mixed results depending on the country and existing pDST and molecular testing practices [7]. In areas where pDST is well-established and widely used for detecting drug resistant TB, tNGS is less likely to be cost effective. However, where pDST access is limited or incomplete, tNGS can be more cost-effective and provide an opportunity for microbiology result guided therapy to a larger proportion of patients with MDR tuberculosis including the compounding benefits derived from reduced transmission of drug resistant if patients receive adequate treatment more promptly.

The clinical utility and cost-effectiveness of direct tNGS highly dependent on specimen bacterial load, as unsuccessful sequencing results may still require follow-up phenotypic or alternative molecular DST testing. Therefore, direct tNGS have greatest utility as a rapid molecular approach for higher bacterial load specimens. Despite this limitation, it could still provide important clinical and public health surveillance value in resource-limited settings, especially those where access to pDST or WGS remains limited. Our panel includes targets associated with bedaquiline resistance and our utilisation of this panel on a pre-XDR sample (by Xpert) highlighted the best use case. As a reflex test after a positive hit on Xpert-MTB/XDR, this tNGS panel can not only uncover resistances not targeted within Xpert-MTB/RIF and Xpert-MTB/XDR, but could also detect mutations associated with bedaquiline resistance, enabling antimicrobial stewardship for this last-line of defence against TB.

Ongoing advancements in sequencing technology will increase the future utility and accessibility of tNGS. Advances in capture probe hybridisation assays may overcome some of the downsides of tNGS by enabling larger regions of the genome to be captured, including important lineage and phylogenetic markers, with comparable sensitivity and turnaround time [29]. The added value of tNGS is highly context-dependent and unlikely to be realised in low-burden settings (like Australia) where routine WGS is well established, facilitating comprehensive drug resistance detection and phylogenetic analysis [30]. Limited evolutionary information is available if lineage and sublineage markers are included in tNGS assay design, but this does not have utility for transmission tracking. Future studies comparing the analytical performance, cost-effectiveness, and implementation of in-house and commercial tNGS assays would provide additional value for clinical and public health applications, and spurn innovation in this important new diagnostic frontier. Further optimisation studies could also evaluate whether reduced-pool multiplex designs achieve comparable amplification efficiency, target coverage, and resistance locus detection performance while reducing workflow complexity and reagent consumption.

Concordance estimates between tNGS, pDST, and WGS-based gDST are limited by small sample size. The small number of drug-resistant isolates, including limited representation of XDR-TB cases, also limited evaluation of assay performance across broader resistance profiles, but sufficed to demonstrate proof-of-principle performance. Additional studies from high TB incidence settings are needed to further assess the clinical utility of tNGS, especially in settings with rising rates of resistance to new BPaL regimens. Concerns about accuracy of ONT calls have been largely addressed by recent improvements in ONT chemistry and basecalling , in addition we ensured high sequencing depth and variability monitoring. Further validation studies are still required to specify diagnostic accuracy, sensitivity, and specificity for resistance detection compared with established culture-based and short-read sequencing methods. Another limitation of this tNGS panel is that it is more cost effective when batched, which may reduce its uptake for laboratories with low sample throughput.

In conclusion, our in-house tNGS assay provided a fully transparent demonstration of this technology with reliable *M. tuberculosis* lineage identification and drug resistance detection directly from respiratory samples with a high bacterial load (Ct value <27). We quantified the limits of detection and demonstrated the thresholds of sequencing depth required for confident drug-resistance prediction. Given the high cost of commercial tNGS assays, more studies from high tuberculosis incidence settings are needed to clarify its clinical and public health value, with an emphasis on cost:benefit analyses, creative cost saving applications and expanded resistance testing in settings with rising rates of resistance to the new BPaL/M regimens.

## Data Availability

Raw, de-identified tNGS data are available at Zenodo (https://zenodo.org/records/22948274).

## Acknowledgements

The authors thank the Sydney Informatics Hub and the University of Sydney’s high-performance computing cluster, Gadi. The first author (XZ) is funded by NHMRC Centre for Research Excellence in Tuberculosis.

## Author contributions

The study was conceptualised by VS, BM, XZ, and CL. Loci selection and amplicons design were performed by XZ. Genomic DNA preparation was carried out by AB and TC. Multiplex PCR and ONT sequencing were conducted by XZ and ES. Bioinformatic analyses were performed by XZ. ES has accessed and verified the underlying data. The first draft of the Methods and Results was written by XZ and ES, and the Introduction and Discussion by CL. Figures were generated by XZ. All authors participated in manuscript revision and approved the final version.

## Data Summary

The authors confirm that all supporting data and Method details have been provided within the article or as supplementary material. Raw, de-identified tNGS data are available at Zenodo (https://zenodo.org/records/22948274).

## Ethical statement

The study was approved by the Western Sydney Local Health District (WSLHD) Human Research Ethics Committee (approval no. 2019/PID14240).

## Supplementary data

>fgd1_3

TTCGCCCGGCTGCGTGAATCGgtggggctaatgcggcagctgtggagcggtgaccgcgtcgactttgacggcgactattaccggctcaagggtgcctcgatctacgacgtgcccgacgggggcgtgcccgtctacatcgccgccggcggcccggcggtggccaagtacgccggccgcgccggtgacggcttcatctgtacgtccggcaagggcgaggagctctacaccgagaagctgatgccggcggtacgagaaggcgccgctgccgctgaccgatccgtcgacggcatcgacaagatgatcgaaatcaagatctcctacgaccccgacccggagctggcattgaacaacacccggttttgggcgccgctgtcgttgacagctgagcagaagcacagcatcgacgacccgatcgagatggagaaggccgccgatgcgctgccaatcgaacagatcgccaagcgctggatcgtggcgtcggaccccgacgaagccgtcgaaaaggtaggtcaatacgtgacatggggcctgaaccacctggtatttcacgcaccaggacatgaccagcgccggtttctggagctcttccagtcggacctggcacccaggttgcggcgacttggctgactcctcggcgatctacctcgccgcaccagaatCGCAGACGGGTAAGTCGACGATT

>atpE

CCAAGCGATGGAGCTCGAAGAGGaacaccactagtaccggatgctggtaacggctaccagagccatcaaggaggataaggaaatggaccccactatcgctgccggcgccctcatcggcggtggactgatcatggccggtggcgccatcggcgccggtatcggtgacggtgtcgccggtaacgcgcttatctccggtgtcgcccggcaacccgaggcgcaagggcggctgttcacaccgttcttcatcaccgtcggtttggttgaggcggcatacttcatcaacctggcgtttatggcgctgttcgtcttcgctacacccgtcaagtaattcgacggcaaatggttgcaataggtagcaatgggtgaagtgagcgcgattgtcctggccgccagtcaggcggcagaggaaggcggCGAGTCCAGCAACTTCCTCATTCCC

>L9_valS

CCTTCGTCCATGGTGAACCGgtcgcggctccagtccaccccgtcaccgagtcggcgcatctggccgccgatggcaccgccagactctcgcttccaatcccacaccttgtccacgaacagctcgcggccgaggtcttctttagtcttgccgtcgaccgccagctgctgctcgaccacgctctgggtggcgatcccggcatggtcggtgcccggctgccagagcacctcatagccctgcatccgcttgcgccgcgtcaaggcgtccatcatggtgtgttccagcgcgtggcccatgtgcaggctgccggtcacgttcggcggcggcagcacgatcgaataggccggcttggtgctggtcgggtccgcggtgaagtagccagcgtccagccacttctgatagatggcgctctccatcgcggccggatcccaCGACTTGGGCAGCATATCGG

>pncA

CCGCCGCCAACAGTTCATcccggttcggcggtgccatcaggagctgcaaaccaactcgacgctggcggtgcgcatctcctccagcgcggcgacggtggtatcggccgacacacccgctgtcaggtccaccagcaccctggtggccaagccattgcgtaccgcgtcctcggccgtctggcgcacacaatgatcggtggcaataccgaccacatcgacctcatcgacgccgcgttgccgcagccaattcagcagtggcgtgccgttctcgtcgactccttcgaagccgctgtacgctccggtgtaggcacccttgtagaacaccgcctcgattgccgacgtgtccagactgggatggaagtccgcgccgggagtaccgctgacgcaatgcggtggccacgacgaggaatagtccggtgtgccggagaagtggtcacccgggtcgatgtggaagtccttggttgccacgacgtgatggtagtccgccgcttcggccaggtagtcgctgatggcgcgggccagcgcggcgccaccggttaccgccagcgagccaccctcgcagaagtcgttctgcacgtcgacgatgatcaacgcccgcatacgtccaccatacgttcgggcgactgcccgggcagtttgcctaccgaCGCGGCAGCCACAGATATAG

>tlyA_3

AACGATCCTCGGGTGGTGGtcctcgagcggaccaacgcacgtggcctcacaccggaggcgatcggcggtcgcgtcgacctggtagtggccgacctgtcgttcatctcgttggctaccgtgttgcccgcgctggttggatgcgcttcgcgcgacgccgatatcgttccactggtgaagccgcagtttgaggtggggaaaggtcaggtcggccccggtggggtggtccatgacccgcagttgcgtgcgcggtcggtgctcgcggtcgcgcggcgggcacaggagctgggctggcacagcgtcggcgtcaaggccagcccgctgccgggcccatcgggcaatgtcgagtacttcctgtggttgcgcacgcagaccgaccgggcattgtcggccaagggattggaggatgcggtgcaccgtgcgattagcgagggcccgtagtgaccgctcatcgCAGTGTTCTGCTGGTCGTCC

>fabG1_inhA_1

AACCCCAGTGCGAAAGTTCCcgccggaaatcgcagccacgttacgctcgtggacataccgatttcggcccggccgcggcgagacgataggttgtcggggtgactgccacagccactgaaggggccaaacccccattcgtatcccgttcagtcctggttaccggaggaaaccgggggatcgggctggcgatcgcacagcggctggctgccgacggccacaaggtggccgtcacccaccgtggatccggagcgccaaaggggctgtttggcgtcgaatgtgacgtcaccgacagcgacgccgtcgatcgcgccttcacggcggtagaagagcaccagggtccggtcgaggtgctggtgtccaacgccggcctatccgcggacgcattcctcatgcggatgaccgaggaaaagttcgagaaggtcatcaacgccAACCTCACCGGGGCGTT

>L4Mcaprae_purM

CTCAGGCGATGGATAAGCACGacacggtgggcctggacctggtggcgatggtggtcgatgacttggtggtttgcggcgccgagccgctgttcctgttggattacatcgccgtcggtcggatcgtgccggagcgactcagcgcgatcgtcgccggtatcgccgatgggtgcatgcgtgccggctgtgcgctgcttggcggcgagaccgcagaacatccgggcctgatcgagcccgatcactacgatatctctgccaccggcgtcggcgtcgtcgaggcggacaatgtgctgggtcccgaccgggtcaaacccggcgacgtcatcatcgcgatgggctcgtcgggtctgcattccaatgggtactcgctggtccgcaaggtgttgctggagatcgaccggatgaatctggccggtcatgtggAGGAGTTCGGTCGCACCTT

>rpsA_3

CGCAGCGAGTTCCTGAATAACTtgcaaaaaggcaccatccgaaagggtgtcgtgtcctcgatcgtcaacttcggcgcgttcgtcgatctcggcggtgtggacggtctggtgcatgtctccgagctatcgtggaagcacatcgaccacccgtccgaggtggtccaggttggtgacgaggtcaccgtcgaggtgctcgacgtcgacatggaccgtgagcgggtttcgttgtcactcaaggcgactcaggaagacccgtggcggcacttcgcccgcactcacgcgatcgggcagatcgtgccgggcaaggtcaccaagttggttccgttcggtgcattcgtccgcgtcgaggagggtatcgagggcctggtgcacatctccgagctggccgagcgtcacgtcgaggtgcccgatcaggtggttgccgtcggcgaCGACGCGATGGTCAAGGTC

>eis

CAGGCGGTCGCTGATTCAGggccgatgaaatcggtgaaactggccgcggccagtaggaacatccccggccagtcgtcctcggtcgggctacacagggtcacagtcacagaatccgactgtggcatatgccgcggccacgtgcacgtgaatattacgacgacagtgtctggcaaaggatcacgcgatgcgggtagccccgccagcgtgacgccactgcgagaatcagcgacgaatttcgccgtgacgttacgctggcggcgacgctcccacgtcgacgcataccccgacggctccggcaccgacgtgcagagcaagtaccggtcccatggcggtcaccatggccggctcacacgccggcagccgctccgccagcgccgccgccacgtcgttcgcagctgccgggtcggcgacgtgatgcaccgcgagagcggcggggcggtcgccgacaagctGGCAAACCCGGTCGATCATC

>gyrB

CACCGAGGTCAAATCGTTTGTGcagaaggtctgtaacgaacagctgacccactggtttgaagccaaccccaccgacgcgaaagtcgttgtgaacaaggctgtgtcctcggcgcaagcccgtatcgcggcacgtaaggcacgagagttggtgcggcgtaagagcgccaccgacatcggtggattgcccggcaagctggccgattgccgttccacggatccgcgcaagtccgaactgtatgtcgtagaaggtgactcggccggcggttctgcaaaaagcggtcgcgattcgatgttccaggcgatacttccgctgcgcggcaagatcatcaatgtggagaaagcgcgcatcgaccgggtgctaaagaacaccgaagttcaggcgatcatcacggcgctgggcaccgggatccacgacgAGTTCGATATCGGCAAGCTGC

>rpsA_2

AGGTCGTTTCCGTCGGTGAcgaggtcgaagccctggtgctcaccaaggaggacaaagagggccggctcatcctctccaagaaacgcgcgcagtacgagcgtgcctggggcaccatcgaggcgctcaaggagaaggacgaggccgtcaagggcacggtcatcgaggtcgtcaagggtggcctgatcctcgacatcgggctgcgcggtttcctgcccgcctcgctggtggagatgcgccgggtgcgcgacctgcagccctacatcggcaaggagatcgaggccaagatcatcgagctggacaagaaccgcaacaacgtggtgctgtcccgtcgcgcctggctggagcagacccagtccgaggtgcgcagcgagttcctgaataacttgcaaaaaggcaccatccgaaagggtgtcgtgtcctcgATCGTCAACTTCGGCGCG

>Rv1979c_4

AAGCCGAGAAACGTCAGCGtcttcacactgaacagttgctcggcgtcggcccaggccttgtcggggaaggccactcgcaacagcgtcgagacgaaaaaagaagccaacaccccccaagcgatggacgcggtaatggcgtgggtgacaccgacatagatgccgatccggcgcccaaatgcggccgttgtgtaggcgtaggaggcaccgtttgttctgacgtaccttgccgccgtcgcgaagacgatcgccacgacacccgcgaaaatgccagctaaaacataggccatcggcgcgaagggtcctgcgagcccgatcacctcacctggagttaggaagataccggcgccgattatcgagttgatcccgagcatgacgacgctgcagaaacccagcttgtggatcgcatatcctctcgtccgcgggccgaccacCGCACCAAGGCTGTCTAGC

>gid_2

AACTTGTCCAACGCGGCCaccgcccgtgacaccgcagcgtcgctgccgcccaattggtcctgcacccaggactcctcggcgcgcccccgcacgatctcaacggccacgcccagatctgtcaccatctctcgaagaaactcggtgcggcgcagtagcggttctaggagaactacctggaggtccggccgcgctatcgccaatggcacgcccggcaacccggctccgctaccgatatccacgacccggtcaccgcgttcgaggagctcaccgatcacggcgcagttcagtagatgccggtcccatagcctaccgacttcgcggggtcccaccagcccccgctccacaccgggtcccgccaacgcttcggcgtaccgccgagcaaggccaagccgcggtccgaagatcgcagacgccgcgggctcgatcggagacattacgcactccgccggctcgtgaggtctgtgtcatgtttcacgtgaaaCATTCTCCGCTCTCGAGACG

>rplC_1

CCCTTGAAGCGCGACGATTtcggccttgaaattgatcgcgacaaggctgtgtgcgggcgacacgcccgagcgcggggccggtggacctacgacaggtaaacagcggcgcagtattcggcgcaacgctagatcggtccagaaggaccgggtcgatcggcgcgccggggagcaccggacccggatacgggctcgagtgggagtgaggtaggagaagcgtggcgggacagaagatccgcatcaggctgaaggcctacgaccatgaggccattgacgcttcggcgcgcaagatcgtcgaaaccgtcgtccgcaccggtgccagcgtcgtagggccggtgccgctaccgactgagaagaacgtgtattgcgtcatccgctcaccgcataagtacaaggactcgcgggagcacttcgagatgcgcacacacaagcggttgatcGACATCATCGATCCCACGCC

>fabG1_inhA_5

CGGATGTGTCCAAGGGCATCcacatctcggcgtattcgtatgcttcgatggccaaggcgctgctgccgatcatgaaccccggaggttccatcgtcggcatggacttcgacccgagccgggcgatgccggcctacaactggatgacggtcgccaagagcgcgttggagtcggtcaacaggttcgtggcgcgcgaggccggcaagtacggtgtgcgttcgaatctcgttgccgcaggccctatccggacgctggcgatgagtgcgatcgtcggcggtgcgctcggcgaggaggccggcgcccagatccagctgctcgaggagggctgggatcagcgcgctccgatcggctggaacatgaaggatgcgacgccggtcgccaagacggtgtgcgcgctgctgtctgactggctgccggcgaccacgggtgaCATCATCTACGCCGACGGC

>embB_1

CCGACGCCGTGGTGATATTCggcttcctgctctggcatgtcatcggcgcgaattcgtcggacgacggctacatcctgggcatggcccgagtcgccgaccacgccggctacatgtccaactatttccgctggttcggcagcccggaggatcccttcggctggtattacaacctgctggcgctgatgacccatgtcagcgacgccagtctgtggatgcgcctgccagacctggccgccgggctagtgtgctggctgctgctgtcgcgtgaggtgctgccccgcctcgggccggcggtggaggccagcaaacccgcctactgggcggcggccatggtcttgctgaccgcgtggatgccgttcaacaacggcctgcggccggagggcatcatcgcgctcggctcgctggtcacctatgtgCTGATCGAGCGGTCCATGC

>rpoB_1

AGGACTTCTCCGGGTCGATGtcgttgtcgttctctgaccctcgtttcgacgatgtcaaggcacccgtcgacgagtgcaaagacaaggacatgacgtacgcggctccactgttcgtcaccgccgagttcatcaacaacaacaccggtgagatcaagagtcagacggtgttcatgggtgacttcccgatgatgaccgagaagggcacgttcatcatcaacgggaccgagcgtgtggtggtcagccagctggtgcggtcgcccggggtgtacttcgacgagaccattgacaagtccaccgacaagacgctgcacagcgtcaaggtgatcccgagccgcggcgcgtggctcgagtttgacgtcgacaagcgcgacaccgtcggcgtgcgcatcgaccgcaaacgccggcaaccggtcaccgtgctgctcaaggcgctgggCTGGACCAGCGAGCAGATTG

>fbiC_1

CGGCGTTAAGAGTCGAACCGataccggccatgtccgcagcggccgccgccagtgcctctggcgccgcgaacacaaacgacatctcgtaccttctcctggttcaccacgcggcggctgtcgccgggggcttgttcagacgctggcctctcacggatggtatcgcgatcggctgtgacctgcgccttactccaccaaaccgttggtgccggacggtcgacggcgtgccgagctcggcctggcgctactgttgcgcttatggcgccaaggttggccagcatctcacctggtggggcgtgcggatgatatcagattgcagggaaggtataccaacgtgccgcagcctgtaggtcggaagtccaccgctctgccgagtcccgttgtaccgccccaggcaaatgcctcagcgttgcggcgggtactgcgacgggcccgAGATGGTGTCACGCTGAACG

>fbiC_2

CTCTGCCGAGTCCCGTTGTAccgccccaggcaaatgcctcagcgttgcggcgggtactgcgacgggcccgagatggtgtcacgctgaacgtggatgaggcggccatagcgatgaccgcacgcggtgacgagctggccgacctgtgcgcgagcgccgcgcgggtgcgcgatgcgggtctcgtgtcggccggccggcacgggcccagcggcaggttggcgatcagctattcgcgcaaggtgtttatcccggtcacccggttatgccgggacaattgccactattgcacgttcgtcaccgtgccgggcaagctacgcgcccaaggttccagcacgtatatggaacccgacgagatcctcgacgttgcccgccgaggtgccgaattcggttgcaaggaagcgctattcactctcggtgaccgtccggaggcgcgttggcgcCAGGCACGCGAATGGCT

>embA

ACTGGTTTAGGGACTGGGGCgcgttacagcggttgacgccttactaccccgacgcccagcccgctgatctgaacctaggaacggtgactcgcagcgggctgtggagtccggcgccgttgcgccgcggctagaagtgccgtggccaccgactcggcgacaacctccgcggccccgcatcctcaccgcccttaaccgcgtcgcctaccatcgagcctcgtgccccacgacggtaatgagcgatctcaccggatcgcacgcctagcagccgtcgtctcgggaatcgcgggtctgctgctgtgcggcatcgttccgctgcttccggtgaaccaaaccaccgcgaccatcttctggccgcagggcagcaccgccgacggcaacatcacccagatcaccgcccctctggtatccggggcgccacgcgcGCTGGACATCTCGATCCCCT

>Rv1979c_2

GGTCAGGTGCAGGTTGTCGaaccgcagcgccaacgggaatgcgagcgccaacgacgccgtaattgcgaaggagaccatcggcacgtcgtattggttcttgcgtgacaagcgtgtcggcagaaccccgctgtccgctaacgcggtccaaagccgcggtgcaccgaacgaggccgcgacattgatgccgaacatcgatatcagggctccgacgacgatgatcgttcggaaggtagcgtttccgatggccgcggccagtttcacggtgtcgtccgacgcggcgatcttgttcgatccgagcagcatcgctaccgttagggtgagcaagtagatcgcgccaaccgagaagatcgcgatcggtatagctctcggcaggttccggtccggcgcgtccatttcttcggcggcgttcgcgatcgattcgaaACCGGTGAATGCGTACAACG

>rrs_2

AGGAAGGTGGGGATGACGTCaagtcatcatgccccttatgtccagggcttcacacatgctacaatggccggtacaaagggctgcgatgccgcgaggttaagcgaatccttaaaagccggtctcagttcggatcggggtctgcaactcgaccccgtgaagtcggagtcgctagtaatcgcagatcagcaacgctgcggtgaatacgttcccgggccttgtacacaccgcccgtcacgtcatgaaagtcggtaacacccgaagccagtggcctaaccctcgggagggagctgtcgaaggtgggatcggcgattgggacgaagtcgtaacaaggtagccgtaccggaaggtgcggctggatcacctcctttctaaggagcaccacgaaaacgccccaactggtggggcgtaggccgtgaggggttcttgtctgtagtgggcgAGAGCCGGGTGCATGACA

>fbiC_8

CTGCGCGATATTCAGGACCGtaccggcggcttcaccgagttcgtcccgttgccgttcgtgcaccagaattcaccgttgtacctggccggtgcggcgcgccccgggcccagccatcgcgacaaccgcgcggtacatgctttggcgcggatcatgttgcacggccgcatctcgcacattcagaccagctgggtgaaacttggagtgcggcgcacccaggtgatgctcgaaggtggcgccaacgacctgggcggcacgctgatggaggagaccatctcgcggatggccggttccgaacacggatcggccaagaccgtcgctgagctggtcgcgatcgccgaaggcatcggccgcccggcgcgccagcgcactaccacatacgccctgcttgcggcctagccccggcgacgatgccgggtcgcgggatgcggcccgttgaggagcggggcaatctggcctagccccggcgacgatgccgggtcgcgggatgcggcccGTTGAGGAGCGGGGCAATC

>ddn_2

GCACCTTCCAGAAGATTCCGGtcgcgctgctgaccaccaccggccgcaagaccggccagccgcgggtcaacccgctctacttcctgcgcgacggtgggcgggtcattgtcgcggcctccaagggcggcgcggagaagaacccgatgtggtacctcaacctcaaggccaaccccaaggttcaggtacagatcaaaaaggaagtgctggaccttaccgcgcgggacgcgaccgacgaggagcgcgccgaatattggccacagttggtcacgatgtacccaagttatcaggactaccagtcctggaccgaccgcacgatcccgatcgtggtttgcgaaccctgaccgttcccaacttcgccgaacgtgaagccagggcgagaaaacggccgaaatctcgccctgagttcacgctcggcgcagatAACTAGGCCCCATAGACCGG

>L8_ilvD

CCGAACGCCATTACCACCGcgatggcgttctcgaacgcctccttggtgaggatgtcgcgggcggtgatgccgcggcgcagcagctcgacgacggcctgaccgctgcgacgcgcgaacccgtcgcgccggcggtcggtcgccggcggtgccgcgctgcccggcaacgacatgccgagcgcctcggcggcgctggccatggtgttagcggtgtacatgccgccgcatgccccttcgccggggcagattgcccgctcgatggcatcgacgtcggcgcgactcatcaaaccgcgagagcacgctccgaccgcctcgaaggcgtcaatgatggtgacgtctcgttcgctaccgtcggagagcttggcccggccgggcaaaatagagcccgcgtagaggaacaccgccgccagatccagtcgtgcggcggccatcagcaTTCCGGGCAGCGATTTGTC

>fbiC_6

ACCGATGTGCTTGCCGCcctgcgatcggcggagcgtgcgcccgccggctgcaccgacggcgagtatctggcgcttgccaccgccgacggtcctgcgctggaagccgttgccgcactggctgattcgttgcgccgcgatgtcgtcggcgacgaggtgacctttgtggtcaaccgtaacatcaacttcaccaacatctgctacaccggttgccggttctgcgcgttcgcccagcgaaagggtgacgccgacgcctactcgctgtcggtcggagaggtcgccgaccgggcatgggaggcccacgtcgccggggccaccgaagtatgcatgcagggcggtatcgatcccgagctaccggtcaccggctacgccgatctggttcgtgccgtcaaggcgcgggtgccctccatgcatgtgcacgcgttttccccgatggagatcgccaacggcgtcacCAAGAGCGGGCTGAGCATT

>katG_8

CCTCGCCAACAGCACAGTCgacatcggcgataaccccgcaagaccggcagacgatgtgatggtggttgtcgccgaccctggactcgtagcgcgcgacggagcccgagggttggatctttcgcaccaagcccgcggcggtcagggcatgcagcacgtcgtacacggcttgccgggatacgtcgggcagcgcaaaacgcacggcaccgaaaatcgtttccgtgtcggcgtgtggatgcgcattcactgcttccaggacggcgacgcgcggtcgggtcacgcgcaggtcggccgtccggagctgttcggcgtagtccggtatagaggacacactagacaatatgactcccttttctggaatcagtcaagactttggctagcgtgacaggcgtctgctaggacccgatcgccccggggccgctggATCGTGGGATGGCGGGT

>Morygis_leuS

CGATGACGGGTTCGAAGTCGacatcgaggtgttcaccacgcggcccgacaccttgttcggcgccacgtatctggtgctggctcccgagcacgacttggtcgacgagttggtcgccgcgtcctggccggctggggtcaaccccttgtggacatacggcggcggcacacctggtgaggccatcgccgcctaccggcgtgcgatcgccgccaaatcagacctcgagcgccaggagagcagggaaaagaccggcgtcttcttgggcagctacgccatcaacccggccaacggtgagccggtgccgatcttcatcgccgactacgtgctggccgggtacggtaccggggcaatcatggcggtgccgggtcatgaccagcgggactgggacttcgctcgggcatttggtctaccgatcgtggaagtaattgCCGGCGGCAATATTTCGGA

>rpsL_2

CAGAACGTGAAAGCGCCCAagatagaaagccggtagatgccaaccatccagcagctggtccgcaagggtcgtcgggacaagatcagtaaggtcaagaccgcggctctgaagggcagcccgcagcgtcgtggtgtatgcacccgcgtgtacaccaccactccgaagaagccgaactcggcgcttcggaaggttgcccgcgtgaagttgacgagtcaggtcgaggtcacggcgtacattcccggcgagggccacaacctgcaggagcactcgatggtgctggtgcgcggcggccgggtgaaggacctgcctggtgtgcgctacaagatcatccgcggttcgctggatacgcagggtgtcaagaaccgcaaacaggcacgcagccgttacggcgctaagaaggagaagggctgatgccacgcaaggggcccgcgcCCAAGCGTCCGTTGGTCAA

>Rv1979c_3

TCGGTATAGCTCTCGGCAGGttccggtccggcgcgtccatttcttcggcggcgttcgcgatcgattcgaaaccggtgaatgcgtacaacgcgacaatcgtggccagcgccatactcgagaacgtgcccttgccaatttcggcgacgccaagcaacgagtacggggtcgcgctgtatgccgaccacgccgttgcgtagttgttcacgtgctgggtggtgatgatccacagcccgccgacaatgaatgccgagagcgcgaatgccttgcctaccgttgacgttccgttggcccacttgatcgcccggttgccgaagaggttgatggccaacagcacgccgataaagccgagaaacgtcagcgtcttcacactgaacagttgctcggcgtcggcccaggccttgtcggggAAGGCCACTCGCAACAGC

>fabG1_inhA_4

GCTCGAACTCGACGTGCAAaacgaggagcacctggccagcttggccggccgggtgaccgaggcgatcggggcgggcaacaagctcgacggggtggtgcattcgattgggttcatgccgcagaccgggatgggcatcaacccgttcttcgacgcgccctacgcggatgtgtccaagggcatccacatctcggcgtattcgtatgcttcgatggccaaggcgctgctgccgatcatgaaccccggaggttccatcgtcggcatggacttcgacccgagccgggcgatgccggcctacaactggatgacggtcgccaagagcgcgttggagtcggtcaacaggttcgtggcgcgcgaggccggcaagtacggtgtgcgttcgaatctcgttgccgcaggccctatccggacgctggcGATGAGTGCGATCGTCGGC

>tlyA_2

GTGGTGACCGACAGTGAACGcgcctgggtatcgcgcggagcgcacaaactagtcggtgcgctggaggcgttcgcgatcgcggtggcgggccggcgctgtctggacgcgggcgcatcgaccggtgggttcaccgaagtactgctggaccgtggtgccgcccacgtggtggccgccgatgtcggatacggccagctggcgtggtcgctgcgcaacgatcctcgggtggtggtcctcgagcggaccaacgcacgtggcctcacaccggaggcgatcggcggtcgcgtcgacctggtagtggccgacctgtcgttcatctcgttggctaccgtgttgcccgcgctggttggatgcgcttcgcgcgacgccgatatcgttccactggtgaagccgcagtttgaggtggggaaaggtcaggtcggccccggtggggtgGTCCATGACCCGCAGTTGC

>katG_4

GGATCAGCTTGTACCAGGCCttggcgaactcgtcggccaattcctcggggtgttccagccagcgacgcgtgatccgctcatagatcggatccacccgcagcgagaggtcagtggccagcatcgtcggggagcgccctggcccgccgaacgggtccgggatggtgccggcaccggcgccgtccttggcggtgtattgccaagcgccagcagggctcttcgtcagctcccactcgtagccgtacaggatctcgaggaaactgttgtcccatttcgtcggggtgttcgtccatacgacctcgatgccgctggtgatcgcgtccttaccggttccggtgccatacgagctcttccagcccaagcccatctgctccagcggagcagcctcgggttcggggccgaccagatcggccgggccggcgccatgggtcttaccgaaAGTGTGACCGCCGACGAT

>embB_2

CAGCAAACCCGCCTACTGGgcggcggccatggtcttgctgaccgcgtggatgccgttcaacaacggcctgcggccggagggcatcatcgcgctcggctcgctggtcacctatgtgctgatcgagcggtccatgcggtacagccggctcacaccggcggcgctggccgtcgttaccgccgcattcacactgggtgtgcagcccaccggcctgatcgcggtggccgcgctggtggccggcggccgcccgatgctgcggatcttggtgcgccgtcatcgcctggtcggcacgttgccgttggtgtcgccgatgctggccgccggcaccgtcatcctgaccgtggtgttcgccgaccagaccctgtcaacggtgttggaagccaccagggttcgcgccaaaatcgggccgagccaggcGTGGTATACCGAGAACCTGCG

>pepQ_1

CGGTGATTGTCCACAGCTGGccgtcgattaccaggaccagcccgttcttgaagtcagcagtggtcgccacgtgggtctcctacagaatggccagttctttggggaaccgggtcaacaattccggggtctgcccggcggtttcaggcattttcggcgtcccgccagccactaccaatgtgtcctcgatgcggacaccgccgcggccgggtaaatagacaccgggctccacggtcaccacggagcccgccagtagtgtaccggcggatgtgaccccgatgcccggcgcttcatgtatctgcaggccaacaccgtgtcccagtccgtgaccgaagtgctcgccgtagccggcgtcggcgatcagctggcgcgctgcagcgtccaccccccgcagctcggcacccggcagcaacgcctgccgaccggcctgttgcgcCTCGGCCACCAGCTGATAGA

>rplC_3

CGAGTACCAGGTTGGGCAAGagttgaccgcggagatcttcgccgatggcagctacgtcgatgtgacgggtacctccaagggcaaaggtttcgccggcaccatgaagcggcacggcttccgcggtcagggcgccagtcacggtgcccaggcggtgcaccgccgtccgggctccatcggcggatgtgccacgccggcgcgggtgttcaagggcacccggatggccgggcggatgggcaatgaccgggtgaccgttcttaaccttttggtgcataaggtcgatgccgagaacggcgtgctgctgatcaagggtgcggttcctggccgcaccggtggactggtcatggtccgcagtgcgatcaaacgaggtgagaagtgatggctgcgcaagagcagaagacactcaaaatcgacgtcaagacgccggcgggCAAGGTCGACGGCGCTATC

>fbiC_4

GTCTGTTGGTCGGCATCGGcgagacgctatccgagcgcgccgatacgttacatgcgattcgcaagtcgcacaaggagttcgggcatatccaagaagtgatcgtgcagaacttccgcgccaaggaacacaccgcgatggccgccttccccgatgccggaatcgaggattacctggcgacggttgcggtggcgcggctggtgctgggcccgggcatgcgcatccaggcgccgccgaacctggtgtctggcgacgaatgccgggcgctggttggcgccggggtcgacgactggggcggtgtctcaccgttgacgcccgaccatgtcaaccccgaacggccctggcccgctttggacgagctggcggcggtcaccgccgaagccggctacgacatggtgcagcggctgaccgcgcaacCCAAATACGTACAGGCGGGC

>L2_aftD

CTGCCCAGCCAATATTCGGGcgctgaaaaccatcccatggccgttactccttggacacggcgttcacaccaactattgcatgcggtcttgaccacgagactctgatgtggcgaccaccgatgccgccaccacggaaaccgaaatcagtgccagcagttgcacactggcccagttcccggcgtatccgtcgaccgaccgccacggatgccgggacagcgcagcgccggccaggatcagcccgcccgcagccagtccgacggtgacccggtcacgtagtcgctcccggcggcgcagtgcataccgcacaccgagggccgtgcccatcaccatgaccccggcgatgctggcgatcaccgccccggccgccagcaccccggccgccgcccaggctccgggcctccagggtggtgtgggccggtcggcgagctgccgtcgcccggtccgcCAGAACGCCAGCAGAGCTAG

>fbiB_3

CTCACCAGTGACGGCTTGCccgccgacgcgatagaacgccgggtggcacgcggccagatcctctatgacgcacccgaagtcgtcataccgatgctggtgcccgacggagcacacagctaccccgatgccgcccgcaccgacgccgagcacaccatgttcacggtcgccgtcggagcggccgtacaagccttgctggtcgcgctggccgtgcgcgggctgggcagttgctggatcggctcgacgatctttgccgctgacctggtccgcgacgagctggacctgccagtcgactgggagccgttgggcgccatcgcgatcggatatgccgacgagccgtccgggttgcgcgacccggtgcctgccgccgatttgctgatcctgaagtgacattcgctctagcgacgataggctacccagacatggcggtcctgcagccgatgcCAACCATCAACCTCCCGACG

>katG_5

CAAGCCCATCTGCTCCAGCggagcagcctcgggttcggggccgaccagatcggccgggccggcgccatgggtcttaccgaaagtgtgaccgccgacgatcagcgccgctgtttcgacgtcgttcatggccatgcgccgaaacgtctcgcgaatgtcgaccgccgcggccatggggtccgggttgccgttcggcccctccgggttcacgtagatcagccccatctgcaccgcggccagcgggttctccagatcccgcttaccgctgtaacgctcatcgccgagccaggtggcttccttgccccaatagacctcatcgggctcccactggtcgacccggccgaagccgaacccgaacgtcttgaagcccatcgattccagcgcgcagttgccggcgaaaacaatcaggtccgcccatgagagcttcttgcCGTACTTCTTCTTGACCGGCC

>gid_1

CCCGCTGGAATGGTTCGATAGttgaagcctggcccgaccttacgagcgcggacggtccagcggccaccgggccccacggagcactcacgccgtccctccactcgccatccgtgccgaccctcgggcgatctgctttccacgtcgtgcgaacaccacggtcgcgggcggacgcaaatagttcgcgccacatgtcaccaccctgacatcaaccgcgcccgatgcgatcatcacacgccggtgctcccgtacttcgtcgtgagcccgctcgcctttgatggcgagcattcgcccgttcggccgtatcaacggcatgctccatttcgtcaacttgtccaacgcggccaccgcccgtgacaccgcagcgtcgctgccgcccaattggtcctgcacccaggactcctcggcgcgcccccgcacgatctcaacggccACGCCCAGATCTGTCACCA

>katG_6

AACCCGAACGTCTTGAAGCCcatcgattccagcgcgcagttgccggcgaaaacaatcaggtccgcccatgagagcttcttgccgtacttcttcttgaccggccacagcagccggcgcgccttgtccaagctggcgttgtcgggccagctgttaagcggcgcgaaccgctgcatgccgcccccggcgccgccgcggccgtcgtggatgcggtaggtgccggcagcgtgccacgccatccggataaacagcggcccgtagtggccgtagtcggcgggccaccacggctgcgaggtggtcatcacttcctcgatgtcccgcgtcagggcgtcaacgtcgatggtcgcgacctccgcggcatagtcgaacgccgcacccatcgggtcagcgacggccgggttttggtgcagtacCTTCAGATTGAGCCGGTTGGG

>fbiC_3

CGGTTGCAAGGAAGCGCTAttcactctcggtgaccgtccggaggcgcgttggcgccaggcacgcgaatggctcggcgaacggggctatgactccacgttgtcctacgtgcgcgcgatggcaatccgtgtgctggagcaaaccgggctgttgccgcacctgaacccgggtgtgatgagctggtcggagatgtcgcggctcaaaccggtggcgccgtcgatgggcatgatgctggagacgacctcgcgacggctgttcgaaaccaaggggctcgcccactacggcagccctgacaaagacccggcggtgcggctgcgtgtcctgaccgacgccggccggttgtccattccgtttaccaccggtctgttggtcggcatcggcgagacgctatccgagcgcgccgatacgttacatgcgattcgcaagtcGCACAAGGAGTTCGGGCAT

>tlyA_1

GTCTCTGGCCGAACTCGAAGgcatcgcacgtcgtctttccgaggcgcacgaggtgttgttggccgccctggagtcggcggagaagggttgagtgcggcgtggcacgacgtgcccgcgttgacgccgagctagtccggcggggcctggcgcgatcacgtcaacaggccgcggagttgatcggcgccggcaaggtgcgcatcgacgggctgccggcggtcaagccggccaccgccgtgtccgacaccaccgcgctgaccgtggtgaccgacagtgaacgcgcctgggtatcgcgcggagcgcacaaactagtcggtgcgctggaggcgttcgcgatcgcggtggcgggccggcgctgtctggacgcgggcgcatcgaccggtgggttcacCGAAGTACTGCTGGACCGTG

>katG_1

CGGCGGGTTGTGGTTGATcggcgggcagggccgatcaacccgaatcagcgcacgtcgaacctgtcgaggttcatcaccttgtcccaggcagcgacgaagtcctgcacgaacttcggctgcgcgtcatcggcgccatagacctcgacaagcgcccgcaactccgagttggacccgaagaccaggtccacgcggctgccggtccacttcaccttgccactgccatccttgccctggtaggtcccgtcatctgctggcgagggctcccaggtgatacccatgtcgagcaggttcacgaagaagtcgttggtcagtgactcggaggcctcggtgaacacgcccagcggtaagcgcttgtagtttgcgccgaggacgcgcaggccacctaccagcaccgtcatctcaggggcactgagcgtaagcaggttcGCCTTGTCGAGCAGCATGT

>panD_2

GCCCATGTCGATCGGTTTGTtgtaagcgtcgacaaacacgatccgcggctggtatgtgcgggcccgggcgtcgtccatcgtcgcgtacgcaatcagaatcaccagatcccccggatgcaccaagtgcgcggcggcaccgttgatgccaatcacaccactgccgcgttcgccggtgatcgcgtaggtgaccagtcgagcaccgttgtcgatatcgacgatggttacctgttcgccttccagcaggtcggcggcgtccatcaagtcggcatcgatggtcaccgagccgacgtagtgcaggtcggcgcaggtcaccgtggcgcggtggatcttcgacttcagcatcgtccgtaacatcagtttctccaatgtgattcgaggattgcccggtatccgtccgggcggtcggtgccggcgaaagttccgatttcaatcGCAATGTTGTCCAGCAGCCT

>fbiB_2

CCAACGACCTGTTCTGGCTCgggaccgccgaagcgctcgagctgggtcgccagcaagcccaactgttgcgcaggtccgttcgccggtttagcaccgatccggtgccgggcgacctcgtcgaggctgcggtcgccgaggccctcaccgcgccagccccacatcacacccggccgacccgattcgtgtggctgcagacaccggccatccgcgcgcggctgctagatcggatgaaagacaagtggcggtctgatctcaccagtgacggcttgcccgccgacgcgatagaacgccgggtggcacgcggccagatcctctatgacgcacccgaagtcgtcataccgatgctggtgcccgacggagcacacagctaccccgatgccgcccgcaccgacgccgagcacaccatgttcacggtcgccgtcggagcggCCGTACAAGCCTTGCTGGTC

>L1_hemL

GGTCTACACCGCATTGGACGccaacgctgaccgcctggccggcctgctctccgaggcactgacggatgccgttgtgccacaccagatttcgcgggcaggcaatatgctcagtgtgttcttcggcgaaacaccggtgaccgacttcgcgtccgcgcgggccagccagacctggcgttatccagcgttctttcatgccatgctggacgccggtgtctacccgccgtgcagtgccttcgaggcatggttcgtctcggccgctttggacgacgcggcgttcggccggatcgccaacgcgctgcccgccgcggcccgagcggcggcccaggaaaggcccgcctgatgcccgaggaaacccaagtccacgtggtgcgccacggtgaggtgcacaaccctaccggcatcctgtacgggcggctgcccgGATTCCACCTGTCCGCAACC

>pepQ_3

GTCTCGAATGACACCGCGTCagcgccatgatcgagcatcagggcctccagctcgcggctcacctgccgttcggttcggcccggccgcaggccgccgcgggccaccaagtcggtcagcgcggcatcggctgcttcgcaggctagtcgcagcagcgccagctcgccggcgtctttaacctcgcgcagtgactccacagttccggatgcccgcaccaactcggtgttcttgccctccagcgcgcccgccaaggcgtccaggccgtccaccgtgaccacgtggctctcgaagcccagctttcccacgccggcctcgccggcccggccggccaggtagcgcccgaccgcgcgctcgatagccacttcgaggtcgggcgcttgcgaggcggcctgagtgcggtaccggccgtcggtggccaacacggcatcgcgctcaTCGGCGAACACCAGCAATG

>fbiB_1

GAATCGACTCGGGCTGGTTCaggcggccgccggcgtggacggatccaacgtcggccggtccgagttagcgctgctgccggtcgatcctgacgccagtgccgcaaccttgcgcgccgggctgcgcgagcggctcggcgtcaccgtcgccgtggtcatcaccgacaccatgggacgcgcctggcgcaacggccagaccgatgccgcagtcggcgctgccggtctggcggtgctgcgcaactatgccggtgtccgcgacccatacggcaatgagttggtggtcaccgaggtcgcagtcgccgacgagatcgccgcggccgccgacttggtcaaaggcaaactgaccgcgacgccggtggcggtggtgcgtgggttcggcgtgtccgacgacggctcgacagcccggcaactgctgcggccgggcgCCAACGACCTGTTCTGGCTC

>fbiD_2

GATGTACTGGCCGACCCGAcacccgaagacgatcccgacccactgaacaccgccatcaccgctgccgaacgcgtggttgccgaaggggcctccaacatcgttgtgctgcaaggcgatttgccggcattacagacacaggaactcgccgaggcaatctcggccgcacgccaccatcggcgcagcttcgtcgccgaccggcttgggaccggcaccgcggtactgtgtgcgttcggcaccgcgctgcacccgcggttcgggccggattcgtccgcgcggcaccgccgttcgggcgctgtcgagctgacaggagcctggccgggcctgcgctgcgatgtcgacacccccgccgacctgacggccgcacgccagctcggggtagggcccgcgaccgcgcgagcggtcgcacatcgttgaccgggacggggcaacgccggcgaggcatccaggGGGTGAACGGCAGACCAAC

>L4L2_Rv2509

CACTGGTCGAGAAGCTGGTGccggacttcctgtggatctcgacggagcacaccgcccgggtatcgctgaatgccttggagcgcaacaagatgcgcgtcgttccgggtctgacgtcaaaggcgatgtcggtggccagccaatacgctccgcgcgccatcgtggcgccaatcgtgggtgccttttacaagaggcttgggggcagctaggcatcacttccggcggcggcgcccggtgccgaagatgctgcgggtgatctcgcgtgcggtggtgttgaggacgctcttgacggtcggattcttgagtatctcctcccacaccgcggggccctgcggctccaccggagcgggcatcggcggaacttcaaaatcgtccggccagggcagcggatcgtactgcccccttggggctggggcctcctgggccggggcctcttgcgccggcgcgagtttggcGCTCAGTATCTCGTGGGCTG

>Rv1979c_5

TGACGACGCTGCAGAAACCcagcttgtggatcgcatatcctctcgtccgcgggccgaccaccgcaccaaggctgtctagcagggaatcctctaacgcaccatagattctctagcgacgattcttgagctcccggcctgtcgatgccggcgctgcaggtgagtcaccgcagtgggcgcaccgaacactcatttccgccgccccaaatccgcgcagtgaccaccgcgcggtcctcgcgagtctaggccagcatcgagtcgatcgcggaacgtgggaccaatacctgggttgggccggctgcttcgggcagcaactcccccgggttgaagaagaaaatcaccccgtcgttcgtgactgcgaagttctgataattcaccgggtccaagccggcattcggcgctatcgatacctgttgtCCGGTCTGCTTGCTCAGTTC

>fabG1_inhA_3

GTGGTCAGCTTCCTGGCTTCcgaggatgcgagctatatctccggtgcggtcatcccggtcgacggcggcatgggtatgggccactgacacaacacaaggacgcacatgacaggactgctggacggcaaacggattctggttagcggaatcatcaccgactcgtcgatcgcgtttcacatcgcacgggtagcccaggagcagggcgcccagctggtgctcaccgggttcgaccggctgcggctgattcagcgcatcaccgaccggctgccggcaaaggccccgctgctcgaactcgacgtgcaaaacgaggagcacctggccagcttggccggccgggtgaccgaggcgatcggggcgggcaacaagctcgacggggtggtgcattcgattgggttcatgccgcagaccgggatgggcATCAACCCGTTCTTCGACGC

>ethA_1

ACATTCGTTCCGGGCGATATCgcctacagcgacgacgacggcatcatcgtcgtctgactatggcctaaaccggcgctaaaccgtcgctaaagctaaacccccaccggggcaggccttttggcgaaccgcagaccctcgtcgtcgatcttgccgcgccggatgagccggatgtcacgtaggtagttctgattcaggcgccacggtgtacgcgaaccctgcttgggcagctcgtccagcgagcgcagcacgtaacctggggtgaactccatgaagggccgctcttcgacatctgagcccggtcgctcgacgaccacggtgtcaaaaccgttgtcgtccatgtaattcaacaagcgacagacaaactccgacaccaggtcggccttcagcgtccaggaggcattggtgtagccaaccgtgtaggccatgttggggatgccggaaagcatcatgcccttgtaggccatcgtcgtggtgatgtccacttGTTGTCCGTCGATAGTCGCC

>ddn_1

TGTCCACCGGCACCATCAtcgagcggatttaacctagctgcggcagggcaccgtgcggcgtgactgcaacatgaagcgaccgatgattagatagcgaggcggacgcgcgcctttggcgacccttggtcgctaggatcagcgtcatgccgaaatcaccgccgcggtttctgaattcgccgctcagcgacttctttatcaagtggatgtcacggattaatacctggatgtaccgccgcaacgacggggagggtctgggcggcaccttccagaagattccggtcgcgctgctgaccaccaccggccgcaagaccggccagccgcgggtcaacccgctctacttcctgcgcgacggtgggcgggtcattgtcgcggcctccaagggcggcgcggagaagaacccgatgtggtacCTCAACCTCAAGGCCAACCC

>fbiA_1

GCAGAGGTTCCTCGAATGGCgccggcgcctcgggaaccaaactcagatgcggtcgcaaaactgccgttgctgatgcggtagccgatccggtagtggtatgcggtgtgcctcccattacaccccgaaggtgttcataggacatgcctccgcctcctcactcgatagatagtgaaatggtttcccactgttttgatgtacagttaacccaattcgaacaagtgatcgaatctcggtctgcgacaccgaaaccggccggccaaccgcgaaatgacactgatgtgattagacacaagttggggacgcgggtcaagtgtgccggcgcatttccatatcatctcgtaataaaatttccgcggttctgttgtggttgggtcccggcgtgtcgagcgtgactcgtaaccaacgtttggtgatgggcgccGGGAGGTACTGTCCTGCGAT

>L7_glmU

ACGTCGGTGAGGTAGAGCTCctgttgggcgttgttggagctcagccggctcagtgcggaccgcagcgcggcgatgtcgaaggcgtagacgccggcgttgacttcgcggatttcccgctgcgatggtgtcgcgtcggtttgctccacgatcgccatgacttcgtgatcctgggtgcgcaggatgcggccgtagccgaagggatcatccagcgtcgtggtcagcaccgtcaccgcagccgacaccgcgcggtgggtggcgatcaagtcggccagcgtgtcggcgtccagcagcggggtatctcccgaggtgaccacgacgttgccggcgtagtcatcgggcagcgcggacagcccgcagagtaccgcatgcccggtccctagcggtcgatcctgcagggcgacgtcgatcgttcggcCTAGGGTGTCGGCGAGTTCA

>ethA_5

CAGGGACGGAATCGGAAACCtagcgtgtacatgtcggagtcggagcgaattccgggataacggaacaaatcccaggtgccgcccatggattcccgcttttccaggatggcgtagctcttggtcgggcaacggtcctgcaggtgccaggccgcgctgacaccggagattccagcgcccacgatgacaacgtcgaggtgctcggtcatggatccacgctatcaacgtaatgtcgaggccgtcaacgagatgtcgacactatcgacacgtagtaagctgccagggtgaccacctccgcggccagtcaggcttcgctgcctaggggccggcgcaccgcgcggccgtccggcgacgatcgtgaactggcgatcctcgccaccgccgagaaccttctcgaggaccgTCCGCTGGCCGATATCTCG

>fbiA_3

AGGAACTGGTGCGCTATGGcgtgcagcccgactggttcgagctcggggaccgcgatctggccacccatctggtgcgcacccagatgctgcaggccggctaccccctgtcacagatcaccgaggccctatgcgatcgctggcaaccgggcgcccgcttgctgcctgccaccgacgaccgttgcgaaacccatgtagtgatcaccgacccggtcgacgaaagccgcaaggcgatccattttcaggagtggtgggtgcgctaccgtgcccaggtgccgacgcacagctttgcttttgtcggcgctgaaaagtccagcgctgcaaccgaagcgatcgccgccctggccgacgccgacatcatcatgctggcgccgtctaatccggtggtcagcatcggcgccatcctggccgtccccgggattcgcgcggcgttgcgGGAAGCAACCGCACCGAT

>rrs_1

GAGAAGAAGCACCGGCCAACtacgtgccagcagccgcggtaatacgtagggtgcgagcgttgtccggaattactgggcgtaaagagctcgtaggtggtttgtcgcgttgttcgtgaaatctcacggcttaactgtgagcgtgcgggcgatacgggcagactagagtactgcaggggagactggaattcctggtgtagcggtggaatgcgcagatatcaggaggaacaccggtggcgaaggcgggtctctgggcagtaactgacgctgaggagcgaaagcgtggggagcgaacaggattagataccctggtagtccacgccgtaaacggtgggtactaggtgtgggtttccttccttgggatccgtgccgtagctaacgcattaagtaccccgcctggggagtacggccgcaaggctaaaactCAAAGGAATTGACGGGGGCC

>ethA_4

TGCCGATCACGACGATGTTCttagcgtcgtagtcgaggtcctcgggccagtgctgcggatggatgatcggcccgacgaaatcctccgagccggcgaatctcggcgagtagccctcgtcgtagttgtagtagccgctgcacagaaagaggaattcgcaggtgagggcgctgagcgtgccgtggctttggatgtgaacggtccagcggttttccgcggtcgaccaatcggcactgatcaccttgtggtggaaccggatatgcctgtcgattccatacatggccgcggtgctcttgacgtactcgaggatgggcttgccgtcggcgatcgcctgccgtccggtccagggacggaatcggaaacctagcgtgtacatgtcggagtcggagcgaattccgggataacggAACAAATCCCAGGTGCCGC

>Rv1979c_1

GTGTTCTCCCGTTCGAGCTCagcgccaacggaatctagcgagtgctcggccgatcgccaacccggcgaatgattcggtagtagtgcagataagccatcgccggtaccacgatgaacgtgatcacgatcaaagcaatcgagaagtagttcggaccaccccgcactagaaagatgcagcggtagtcgtaggacactgccagcccaaccgagaccacgatcgcaacaagcggtaacaccttgtcggtgaacgcatttcgccgcacagcagcatgttctactgcctgagacctcgccaatgcgatgagagcgatcggcacgatgatgaactggacgaatcgggcgatcaccgccaggccggtcaggtgcaggttgtcgaaccgcagcgccaacgggaatgcgagcgccaacgacgccgtaattgcgaaggAGACCATCGGCACGTCGTA

>fbiAB_2

TGCTGATGACCGACCCGAAcgcgacggctgagatggttcgcgccgggtgcgaccttgcgggagtggtagcttgaccggccccgaacatggctccgcctcgaccatcgagatcctgcccgtcatcgggctgcccgaattccgtcccggcgacgatctgagcgccgccgtcgccgcggcggcaccgtggctacgcgacggtgacgtcgtggtggttaccagcaaggtggtgtccaaatgcgagggccggctggttccggctcccgaagaccccgagcaaagagaccgattgcgccgcaagctgatcgaggatgaggcagtgcgcgtgttggcgcgcaaggaccgcacgttgatcaccgagaatcgactcgggctggttcaggcggccgccggcgtggacggatccaacgtcggccgGTCCGAGTTAGCGCTGCTG

>rpsA_1

TTTTCCGCCCTGAGTTCACGctcggcgcaatcgggaccgagtttgtccagcgtgtacccgtcgagtagcctcgtcaggtaccaatctgtccctacgacccaaccctgtccggagcaacccaacaatatgccgagtcccaccgtcacctcgccgcaagtagccgtcaacgacataggctctagcgaggactttctcgccgcaatagacaaaacgatcaagtacttcaacgatggcgacatcgtcgaaggcaccatcgtcaaagtggaccgggacgaggtgctcctcgacatcggctacaagaccgaaggcgtgatccccgcccgcgaactgtccatcaagcacgacgtcgaccccaacgaggtcgtttccgtcggtgacgaggtcgaagccctggtgctcaccaaggaggacaaagagggCCGGCTCATCCTCTCCAAGA

>katG_3

GCCGCGGAGTTGAATGACTCctggatctcttccagggtgcgaatgaccttgcgcagatccccgtcggggtcgttgacctcccacccgacttgtggctgcaggcggatgcgaccaccgttggcgccgccgcgcttgtcgctaccacggaacgacgacgccgccgcccatgcggtcgaaactagctgtgagacagtcaatcccgatgcccggatctggctcttaaggctggcaatctcggcttcgccgacgaggtcgtggctgaccgcagggaccggatcctgccacagcagggtctgcttggggaccagcggcccaaggtatctcgcaacgggacccatgtctcggtggatcagcttgtaccaggccttggcgaactcgtcggccaattcctcggggtgttccagccagcgacgcgtgatccGCTCATAGATCGGATCCACCC

>katG_7

GCATAGTCGAACGCCGCAcccatcgggtcagcgacggccgggttttggtgcagtaccttcagattgagccggttgggccaccagtcctggtttccgccgccctcgacggggtatttcatatgacccacgacgggacagccgttgctagcggctccggtggtggtttctgtaatgggtgggtgttgctcgggcacagcattccttccaggagttggtgttatcgggctgtgatcacggatgtgatcgcgaagtgtcggatatcgaacaatcaggacatagaccccagtagatgacctccgcctcgtccaacaggaagccgttatggtccgaggccgtcagacagggtgcctcgccaacagcacagtcgacatcggcgataaccccgcaagaccggcagacgatgtGATGGTGGTTGTCGCCGAC

>gyrA

CGCAGCTTTATCACCCGCAacgccaaggatgttcggttcctggatgtctaacgcaaccctgcgttcgattgcaaacgaggaatagatgacagacacgacgttgccgcctgacgactcgctcgaccggatcgaaccggttgacatcgagcaggagatgcagcgcagctacatcgactatgcgatgagcgtgatcgtcggccgcgcgctgccggaggtgcgcgacgggctcaagcccgtgcatcgccgggtgctctatgcaatgttcgattccggcttccgcccggaccgcagccacgccaagtcggcccggtcggttgccgagaccatgggcaactaccacccgcacggcgacgcgtcgatctacgacagcctggtgcgcatggcccagccctggtcgctgcgctacccgctggtggacggccagggcaacttcggcTCGCCAGGCAATGACCCA

>pepQ_2

GTAGTGTACCGGCGGATGTGaccccgatgcccggcgcttcatgtatctgcaggccaacaccgtgtcccagtccgtgaccgaagtgctcgccgtagccggcgtcggcgatcagctggcgcgctgcagcgtccaccccccgcagctcggcacccggcagcaacgcctgccgaccggcctgttgcgcctcggccaccagctgatagatctctagctgccagtcggcggccttgcccaacacgaaggtgcgggtcatatcggagtggtacccggcgaccagggcgccgaagtcgatcttcacgaaatcgccgacctgcagcaccgcgtcggtcggccggtggtgcgggatcgccgaattggccccggcagccacgatcgtctcgaatgacaccgcgtcagcgccatgatcgagcatcagggcctccagctcgcggcTCACCTGCCGTTCGGTTC

>hsp65

ACCAACGATGGTGTGTCCATcgccaaggagatcgagctggaggatccgtacgagaagatcggcgccgagctggtcaaagaggtagccaagaagaccgatgacgtcgccggtgacggcaccacgacggccaccgtgctggcccaggcgttggttcgcgagggcctgcgcaacgtcgcggccggcgccaacccgctcggtctcaaacgcggcatcgaaaaggccgtggagaaggtcaccgagaccctgctcaagggcgccaaggaggtcgagaccaaggagcagattgcggccaccgcagcgatttcggcgggtgaccagtccatcggtgacctgatcgccgaggcgatggacaaggtgggcaacgagggcgtcatcaccgtcgaggagtccaacacctttgggctgcagctcgagctcaccgAGGGTATGCGGTTCGACAAG

>panD_3

CGAGTTCGCTGTCGGTGACcgccaatagcagcagctcagcgctggccgcgacgtccagcggtggcagcaccggggtatcaggcagccggcgctgcgcgcgccgccgggacgcatgagagatggcgctgcacgccaccacaacatggtcggcgcgctgcagcgcgacccctagcgcggtgccgacccggccagccgagatgatccccaccttgagcctggccggacgcaaaccgtcgaaccgctccatagcagacggcctcacaggtttcttggttcgttccagtcccatgcccgggtaccggacggtcaccaagactgtagtcgatttgcacgtcaagacccacccggggcactgctgatttggtcactacaccaacagtgtcggttgccggcggcaatcgggcgGGTACACCCTGGCACAAGC

>L6_serC

GATCACATCGACTGACGGGTCggtttgcggctccggagcactgccgggatccgacgtgatgatgatcggctcgccgacgaacgggttcttggaaacggcggaagcgaacttcgcgctgaactcgccgtaagtcaagtgcagtgagcgtttgtcaatcagcccgaaggcggccgcatcccagaacgccgtggcaccaccattgcccagtatcacctcatagccgtccggcaacgagaacagctcggccaggcctgaccgaaccctgcccaccagattcttgaccggcgcctgtcggtgcgacgtgccgaacaatgccgctgcggtggtggtcagcgtttgcagttgctcaagccggaccttcgacgggcccgacccaaagcggccgtcgcggggtttgatggcggtgggaatttccaggtggggggtgagctggtcgGCCATGCCATCAGGGTAGTG

>L5_hisD

GCTGCTCTGGGAGAATGGTCtacgtgaccgcgccgccgcccgtgcttacccgtatcgacttgcggggagccgagttgacagctgccgagctgcgggccgctctgccacgcggcggcgccgatgtggaagccgtgctgccgacggtacggcccattgtggcggccgtcgccgagcgcggggccgaggccgcgctggacttcggcgcatcgttcgacggtgtgcggccccatgccatccgggtgccagacgcagcgctggacgcggcgctggccggactggactgcgacgtctgcgaagcgttgcaggtgatggtcgagcggacccgcgccgtgcactccgggcagcgtcgcaccgacgtcacaaccacactgggcccgggcgcgacggtcaccgagcggtgggttccggtcgagcgggtaggcctgtacgtgccggggGGCAATGCGGTGTACCCAT

>fbiC_7

GGTGCCCTCCATGCATGTGcacgcgttttccccgatggagatcgccaacggcgtcaccaagagcgggctgagcattcgcgagtggctgatcggcctgcgcgaggccgggctggataccatcccgggtaccgccgcggaaatcctggacgacgaggttcgctgggtgctgaccaagggcaagctgccgacgtcattgtggatcgaaatcgtgacgaccgcccacgaggtgggtctgcggtcatcatcgacgatgatgtacgggcatgtggacagtccacggcactgggtcgcccatcttaacgtgctgcgcgatattcaggaccgtaccggcggcttcaccgagttcgtcccgttgccgttcgtgcaccagaattcaccgttgtacctggccggtgcggcgcgccccgggcccagccatcgcgacaaccgcgcgGTACATGCTTTGGCGCGGA

>Rv0678

CTGGTGACGCATACCGAACGtcacagatttcagagtacagtgaaacttgtgagcgtcaacgacggggtcgatcagatgggcgccgagcccgacatcatggaattcgtcgaacagatgggcggctatttcgagtccaggagtttgactcggttggcgggtcgattgttgggctggctgctggtgtgtgatcccgagcggcagtcctcggaggaactggcgacggcgctggcggccagcagcggggggatcagcaccaatgcccggatgctgatccaatttgggttcattgagcggctcgcggtcgccggggatcggcgcacctatttccggttgcggcccaacgctttcgcggctggcgagcgtgaacgcatccgggcaatggccgaactgcaggacctggctgacgtggggctgagggcgctgggcgacgccccgccgcagcgaagccgacggctgcgggagatgcgggatctgttggcatatatggagaacgtcgtctccgacgccctggggcgatacagccagcgaaccggagaggacgactgatgagcaacctcgcAATCTGACCGAGGTGGCGA

>rrl_1

GCAAGGGTGAAGCGGAGAATttaagccccagtaaacggcggtggtaactataaccatcctaaggtagcgaaattccttgtcgggtaagttccgacctgcacgaatggcgtaacgacttctcaactgtctcaaccatagactcggcgaaattgcactacgagtaaagatgctcgttacgcgcggcaggacgaaaagaccccgggaccttcactacaacttggtattgatgttcggtacggtttgtgtaggataggtgggagactgtgaaacctcgacgccagttggggcggagtcgttgttgaaataccactctgatcgtattgggcatctaacctcgaaccctgaatcgggtttagggacagtgcctggcgggtagtttaactggggcggttgcctcctaaaatgtaacggaggcgcccaaaggttccctcaacctggacgGCAATCAGGTGGCGAGTGT

>rplC_2

GACATCATCGATCCCACGCCgaagaccgttgacgcgctcatgcgcatcgaccttccggccagcgtcgacgtcaacatccagtaggagattggacagagcaatggcacgaaagggcattctcggtaccaagctgggtatgacgcaggtattcgacgaaagcaacagagtagtaccggtgaccgtggtcaaggccgggcccaacgtggtaacccgcatccgcacgcccgaacgcgacggttatagcgccgtgcagctggcctatggcgagatcagcccacgcaaggtcaacaagccgctgacaggtcagtacaccgccgccggcgtcaacccacgccgatacctggcggagctgcggctggacgactcggatgccgcgaccgagtaccaggttgggcaagagttgaccgcggagatcttcgccgatggcagctacgTCGATGTGACGGGTACCTCC

>Rv1979c_6

GCGCTGCAGGTGAGTCAccgcagtgggcgcaccgaacactcatttccgccgccccaaatccgcgcagtgaccaccgcgcggtcctcgcgagtctaggccagcatcgagtcgatcgcggaacgtgggaccaatacctgggttgggccggctgcttcgggcagcaactcccccgggttgaagaagaaaatcaccccgtcgttcgtgactgcgaagttctgataattcaccgggtccaagccggcattcggcgctatcgatacctgttgtccggtctgcttgctcagttcaccttgcacaatggggaagacgactggcagcggatcggtgtcagcctgccacagcgtgtcataggtgattggcttgcgataggcctggtcccaatcgaaggccttgtacgtggtcgttgggtgcgtgccgcCGGCGTTCTGGTAGACCTTG

>fgd1_1

TGAACAAGCTCCGCCATGCccgcgagtctaggagcgagcgcgagcgcggcaagccgggtgccgcgggtcgcgaccatgggatatggagcgatcgcgagcgcggcgaagccgggcgtggcgggtcgcgtttatggcataggagtagaaagaactggtggctgaactgaagctaggttacaaagcatcggccgaacaattcgcaccgcgcgagctcgtcgaactagccgtcgccgccgaagcccacggcatggacagcgcgaccgtcagcgaccattttcagccttggcgccaccagggcggccatgccccgttctcgctgtcctggatgaccgctgtcggcgaacgtaccaaccggctgctgctgggcacttcggtgctgacccccaccttccgctacaaccccgccgtcatcgctcaggcTTTCGCCACCATGGGATGC

>fgd1_2

TACCAACCGGCTGCTGCTgggcacttcggtgctgacccccaccttccgctacaaccccgccgtcatcgctcaggctttcgccaccatgggatgcctgtacccgaaccgtgttttccttggcgtgggcaccggtgaggcgctgaacgaaatcgccaccggatacgagggcgcctggccggagttcaaggagcggttcgcccggctgcgtgaatcggtggggctaatgcggcagctgtggagcggtgaccgcgtcgactttgacggcgactattaccggctcaagggtgcctcgatctacgacgtgcccgacgggggcgtgcccgtctacatcgccgccggcggcccggcggtggccaagtacgccggccgcgccggtgacggcttcatctgtacgtccggcaagggcgaggagctctacaccgagaagctgatgccggcggTACGAGAAGGCGCCGCT

>rrl_2

GCGGGTAGTTTAACTGGGGCggttgcctcctaaaatgtaacggaggcgcccaaaggttccctcaacctggacggcaatcaggtggcgagtgtaaatgcacaagggagcttgactgcgagacttacaagtcaagcagggacgaaagtcgggattagtgatccggcacccccgagtggaaggggtgtcgctcaacggataaaaggtaccccggggataacaggctgatcttccccaagagtccatatcgacgggatggtttggcacctcgatgtcggctcgtcgcatcctggggctggagcaggtcccaagggttgggctgttcgcccattaaagcggcacgcgagctgggtttagaacgtcgtgagacagttcggtctctatccgccgcgcgcgtcagaaacttgaggaaacctgtccctagtacgagaggaccgGGACGGACGAACCTCTGGT

>rpoB_2

TACGGTCGGCGAGCTGATccaaaaccagatccgggtcggcatgtcgcggatggagcgggtggtccgggagcggatgaccacccaggacgtggaggcgatcacaccgcagacgttgatcaacatccggccggtggtcgccgcgatcaaggagttcttcggcaccagccagctgagccaattcatggaccagaacaacccgctgtcggggttgacccacaagcgccgactgtcggcgctggggcccggcggtctgtcacgtgagcgtgccgggctggaggtccgcgacgtgcacccgtcgcactacggccggatgtgcccgatcgaaacccctgaggggcccaacatcggtctgatcggctcgctgtcggtgtacgcgcgggtcaacccgttcgggttcatcgaaacgccgtaccgcaaggtggtcgacggCGTGGTTAGCGACGAGATCG

>fbiA_2

TTCCGCGGTTCTGTTGTGGttgggtcccggcgtgtcgagcgtgactcgtaaccaacgtttggtgatgggcgccgggaggtactgtcctgcgatgtgaaggtcaccgttctggccggtggagtcggcggcgcccgcttcctgctcggggtccagcagctgctcggcctgggccagtttgctgccaattctgcccactcggacgccgaccaccaactgagcgctgtcgtcaacgtcggcgacgacgcctggatccacgggctgcgtgtctgcccggatctggacacctgcatgtataccctgggcggcggggtggacccccagcgcggctggggccagcgtgacgaaacttggcacgccatgcaggaactggtgcgctatggcgtgcagcccgactggttcgagctcggggaccgCGATCTGGCCACCCATCTG

>fabG1_inhA_2

TGTCCAACGCCGGCCTATccgcggacgcattcctcatgcggatgaccgaggaaaagttcgagaaggtcatcaacgccaacctcaccggggcgttccgggtggctcaacgggcatcgcgcagcatgcagcgcaacaaattcggtcgaatgatattcataggttcggtctccggcagctggggcatcggcaaccaggccaactacgcagcctccaaggccggagtgattggcatggcccgctcgatcgcccgcgagctgtcgaaggcaaacgtgaccgcgaatgtggtggccccgggctacatcgacaccgatatgacccgcgcgctggatgagcggattcagcagggggcgctgcaatttatcccagcgaagcgggtcggcacccccgccgaggtcgccggggtggtcagcttcctggcttccgaggatGCGAGCTATATCTCCGGTGC

>L3_eccA3

GAGGAGTTTGCCAAGGCCAAtcacgtggtgtccggtatcaccgagcgccgcgccggctggcgtgccgcccgttggctcgccgtggtcatcaactaccgcgccgagcgctggtcggatgtcgtgaagctgctcactccgatggttaatgatcccgacctcgacgaggccttttcgcacgcggccaagatcaccctgggcaccgcactggcccgactgggcatgtttgccccggcgctgtcttatctggaggaacccgacggtcctgtcgcggtcgctgctgtcgacggtgcactggccaaagcgctggtgctgcgcgcgcatgtggatgaggagtcggccagcgaagtgctgcaggacttgtatgcggctcaccccgaaaacgaacaggtcgagcaggcgctgtcggataccAGCTTCGGGATCGTCACCA

>rpsL_1

GTGGTACCGCGTCAACGTAGcgccctgcttcggccgcaacgcccgctttgacctgccagactggcggcgggtattgtggttgctcgtgcctggcggcttacgcttgatgtaggggcgtggatgccgggccaattcgcatgtccgcgatgcctcggatgagacgaatcgagtttgaggcaagctatgcgacacacccggccgcgggtaaccgtggcggggcatggccgacaaacagaacgtgaaagcgcccaagatagaaagccggtagatgccaaccatccagcagctggtccgcaagggtcgtcgggacaagatcagtaaggtcaagaccgcggctctgaagggcagcccgcagcgtcgtggtgtatgcacccgcgtgtacaccaccactccgaagaagccgaactcggcgcttcggaaggttgcccgcgtgaagtTGACGAGTCAGGTCGAGGTC

>ahpC_1

GCACTGCTGAACCACTGCTttgccgccaccgcggcgaacgcgcgaagcccggccacggccggctagcacctcttggcggcgatgccgataaatatggtgtgatatatcacctttgcctgacagcgacttcacggcacgatggaatgtcgcaaccaaatgcattgtccgctttgatgatgaggagagtcatgccactgctaaccattggcgatcaattccccgcctaccagctcaccgctctcatcggcggtgacctgtccaaggtcgacgccaagcagcccggcgactacttcaccactatcaccagtgacgaacacccaggcaagtggcgggtggtgttcttttggccgaaagacttcacgttcgtgtgccctaccgagatcgcggcgttcagcaagctcaatgacgagttcgaggaccgcgacgcccagatCCTGGGGGTTTCGATTGACAG

>fbiD_1

CGCGACGGTGCTTGCTATTccggccatccacacacactaatctgcgccgcggttgccgtcgggaccgtgcctgggccccggccacgaccgtggcggcaatgccgtcgaagtgtgccgcgtggatcgacgctggcaggatgacttcatgagcggcacaccggacgacggcgatatcggcttgatcatcgccgtcaagcgcttggccgcggccaaaaccaggctggccccggtgttctcggcgcagactcgcgagaacgtggtgctggccatgctcgtcgacacgttgaccgccgcggcgggtgtcggttcactgcgctcgatcactgttatcacccccgacgaagccgcggcggctgcggcggccgggctgggcgccgatgtactggccgacccgacacccgaagacgatcccgacccactgaacaccgccatcaccgctgccGAACGCGTGGTTGCCGAA

>ahpC_2

TCGTGTGCCCTACCGAGATCgcggcgttcagcaagctcaatgacgagttcgaggaccgcgacgcccagatcctgggggtttcgattgacagcgaattcgcgcatttccagtggcgtgcacagcacaacgacctcaaaacgttacccttcccgatgctctccgacatcaagcgcgaactcagccaagccgcaggtgtcctcaacgccgacggtgtggccgaccgcgtgacctttatcgtcgaccccaacaacgagatccagttcgtctcggccaccgccggttcggtgggacgcaacgtcgatgaggtactgcgagtgctcgacgccctccagtccgacgagctgtgcgcatgcaactggcgcaagggcgacccgacgctagacgctggcgaactcctcaaggcttcggcctaaccgggatctggttggccgggaatcaatgagtaTAGAAAAGCTCAAGGCCGCG

>ethA_3

TGTAGTGCGGGCCGAAGTgctttcgcacgtcgtacccctcgggtagctggcgctggatcaggctcaggaacatcttccgcatgcgccgtggccacttctggcaggcgctgtacacggccgcctggcgcagcacgttcttccaccgtaccgcggtgtaggccatggtctccggcagccagcggttgagcttctcggcgatgccgtcccggtctggctgcgacacgatgtaggtgggtgagcgctgcagcatcgtgacgtgcttggcgcccgagtccgccagcgccggcacgagcgtgaccgccgttgcgccactgccgatcacgacgatgttcttagcgtcgtagtcgaggtcctcgggccagtgctgcggatggatgatcggcccgacgaaatcctccgagccggcgaatctcggcgagtagccctCGTCGTAGTTGTAGTAGCCGC

>katG_2

GTCAGTGACTCGGAGGCCTcggtgaacacgcccagcggtaagcgcttgtagtttgcgccgaggacgcgcaggccacctaccagcaccgtcatctcaggggcactgagcgtaagcaggttcgccttgtcgagcagcatgtactcggccggcaacgggttgccctttccgaggtagtttcggaagccatctgccttgggctccagcacggcaaaggattccacgtcggtttgttcctgcgacgcatccgtgcggcccggggtgaagggcaccgtgatgttgtggccagccgcctttgctgctttctctatggcggcacagccaccgagcacgacgaggtcggcgaaggacactttgatgttccccggcgccgcggagttgaatgactcctggatctcttccagggtgcgaatgaccttgcgcagatccccgtcggggtcgtTGACCTCCCACCCGACTTG

>ethA_2

GTGTCAAAACCGTTGTCGTCCatgtaattcaacaagcgacagacaaactccgacaccaggtcggccttcagcgtccaggaggcattggtgtagccaaccgtgtaggccatgttggggatgccggaaagcatcatgcccttgtaggccatcgtcgtggtgatgtccacttgttgtccgtcgatagtcgccgtcgccccaccaaaaagctgcaggttcaaccccgttgcggtaatgatgatgtcagccggcagttcgcgacctgagttcagccggattccggtcgcggtgaaccgttcaatggtgtcggtcaccacctcgaccttcccgtgacgaatggcccggaacaggtcgccgttgggcaccaagcacaatcgctggtcccaggggttgtagtgcgggccgaagtgctttcgcacgtcgtacccctcgggtagctggcGCTGGATCAGGCTCAGGAAC

>panD_1

TGAGCCGCTCGGAATCCTcgccgatcagcccgtcgatcgtcagtgccagttcgtcggcggtgacttcggattcggtgcgtatccgccactgctgcacgacctttgcgtgctctttcattccggacagcaggcccacaacggtgtgggtgttgcggacgtcaatcgccagcagcacggctatcccacaccgagccgggggtctagcagctcgcccgcgttttcgggcacaaatgccggatcgtggcccatgtcgatcggtttgttgtaagcgtcgacaaacacgatccgcggctggtatgtgcgggcccgggcgtcgtccatcgtcgcgtacgcaatcagaatcaccagatcccccggatgcaccaagtgcgcggcggcaccgttgatgccaatcacaccactgccgcgttcgccggtgatcgcgTAGGTGACCAGTCGAGCACC

>L4Mbovis_dnaB

TTTCCTGGATCACACACGCCccaaagttgggcggccacgattcgtggcggctgcacattcatggcgcgaaggatcaggtcaggttccttcgtcacgtcggcgttcacggcgccgaagcggtggcggcccaagagatgctgcgtcagctcaaaggaccggttcgcaacccgaacctggacagcgcgccgaaaaaagtatgggcgcaagtccgcaaccgactgtccgccaaacagatgatggacatccagctccacgaaccgacgatgtggaagcattccccgagccggtcaaggccgcatcgcgcggaggcgcggatcgaagatcgagcgatccatgagctggcgagaggcgacgcgtactgggacaccgtcgtggagatcaccagcattggagatcaacatgttttcgatgggactgtaagcgGCACACACAATTTCGTCGCC

>rpsA_4

TGGTGCACATCTCCGAGCTggccgagcgtcacgtcgaggtgcccgatcaggtggttgccgtcggcgacgacgcgatggtcaaggtcatcgacatcgacctggagcgccgtcggatctcgttgtcgctcaagcaagccaatgaggactacaccgaggagttcgacccggcgaagtacggcatggccgacagttacgacgagcagggcaactacatcttccccgagggcttcgatgccgaaaccaacgaatggcttgagggattcgaaaagcagcgcgccgaatgggaagctcggtacgccgaggccgagcgccggcacaagatgcacaccgcgcagatggagaagttcgccgccgccgaggcggctggacgcggcgcggacgatcagtcgtcggccagtagcgcaccgtcggaaaagaccgcgggTGGATCACTGGCCAGCGA

>fbiC_5

TTTGGACGAGCTGGCGGcggtcaccgccgaagccggctacgacatggtgcagcggctgaccgcgcaacccaaatacgtacaggcgggcgcggcgtggatcgacccgcgggtgcggggacatgtggtggcgctggcggatccggcgaccggcctggcccgcgacgtcaacccggtgggcatgccgtggcaggagcccgacgacgtggcgtcctggggccgggtcgatctgggcgcagcgatcgacactcagggccgcaataccgcagtgcgcagcgacctggccagcgccttcggtgactgggaatcgatccgcgagcaggtgcacgagctggcggtccgcgctccggaacgcattgacaccgatgtgcttgccgccctgcgatcggcggagcgtgcgcccgccggctgcaccgacggcgagtatctggcgcttgccaccgccgACGGTCCTGCGCTGGAA

>pepQ_4

TCGGTGTTCTTGCCCTCCAgcgcgcccgccaaggcgtccaggccgtccaccgtgaccacgtggctctcgaagcccagctttcccacgccggcctcgccggcccggccggccaggtagcgcccgaccgcgcgctcgatagccacttcgaggtcgggcgcttgcgaggcggcctgagtgcggtaccggccgtcggtggccaacacggcatcgcgctcatcggcgaacaccagcaatgcgccgttggacccgctgaagcctgatagatatcgcacgtttatcaggtcgctgatcagcatcgcatccaacccggaggcagcgatttgtgctttcagcttgtctcgacgctgggaatgtgtcacgacccttgacggtactcgctacgctgaatgcccatgactaactggatgctgcgcgggttggcgttcgccgccgcgatggTGGTTCTCCGCCTGTTCCA

>fbiAB_1

CGTCTAATCCGGTGGTCAGCatcggcgccatcctggccgtccccgggattcgcgcggcgttgcgggaagcaaccgcaccgatcgtcggctactcgccgatcatcggcgaaaagccgttgcgcggcatggccgatacgtgcctttcggttatcggggtggattccaccgcggccgctgtgggccggcactacggcgcgcggtgcgccaccgggatactggactgctggctggtgcacgacggcgaccacgctgagattgacggggtgacggtgcggtcggtgccgctgctgatgaccgacccgaacgcgacggctgagatggttcgcgccgggtgcgaccttgcgggagtggtagcttgaccggccccgaacatggctccgcctcgaccatcgagatcctgcccgtcatcgggctgcccgaattccgTCCCGGCGACGATCTGA

---

**Figure S1:**
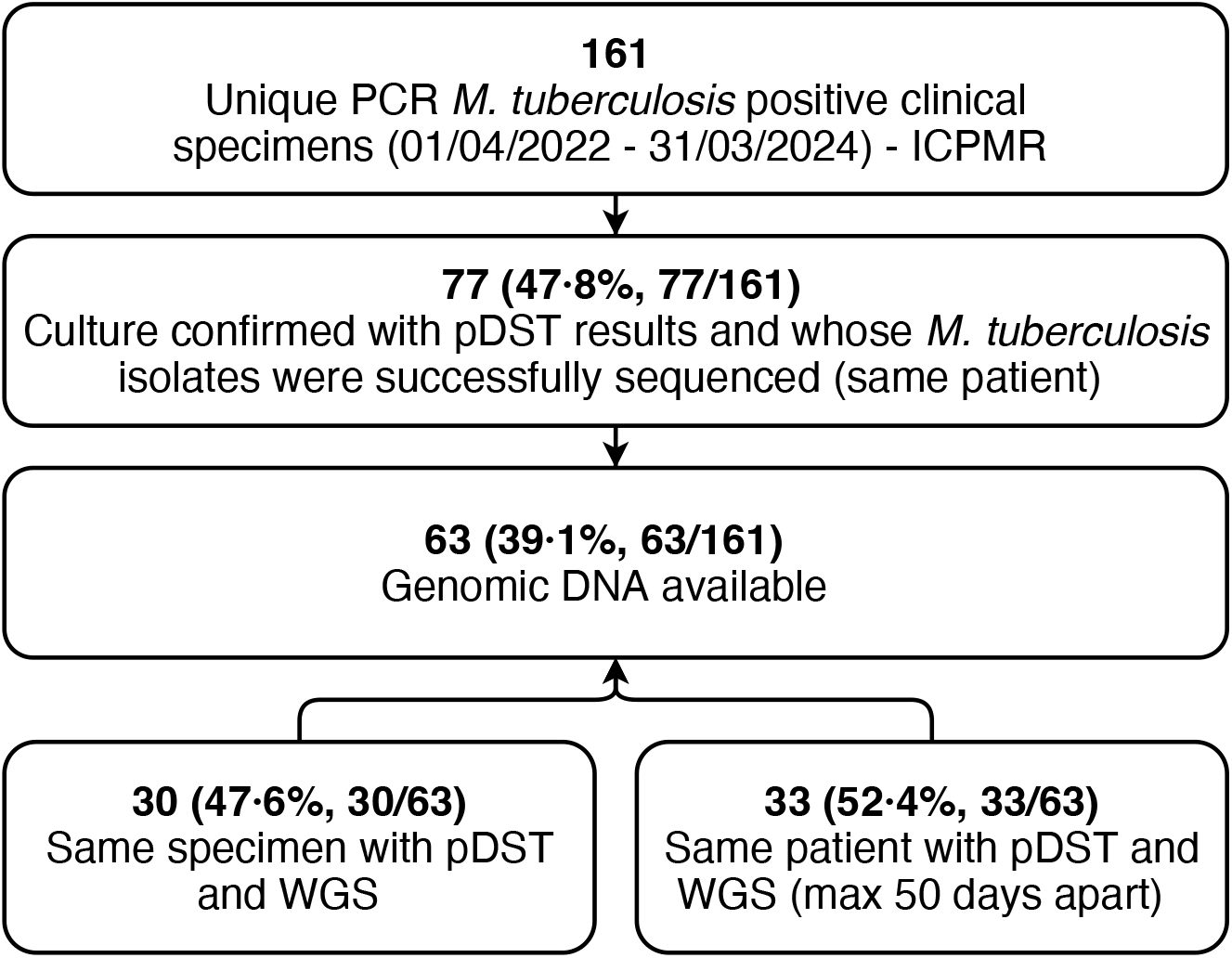
Selection of *M. tuberculosis* clinical specimens. ICPMR: Institute of Clinical Pathology and Medical Research; PCR: Polymerase Chain Reaction; pDST: phenotypic drug susceptibility testing; tNGS: Amplification-based targeted next generation sequencing; WGS: Whole genome sequencing. Genomic DNA extracts from 63 selected *M. tuberculosis* clinical specimens were used for a proof-of-concept evaluation of the tNGS assay, while genomic DNA from the reference strain *M. tuberculosis* H37Rv was used for initial evaluation.

**Figure S2:**
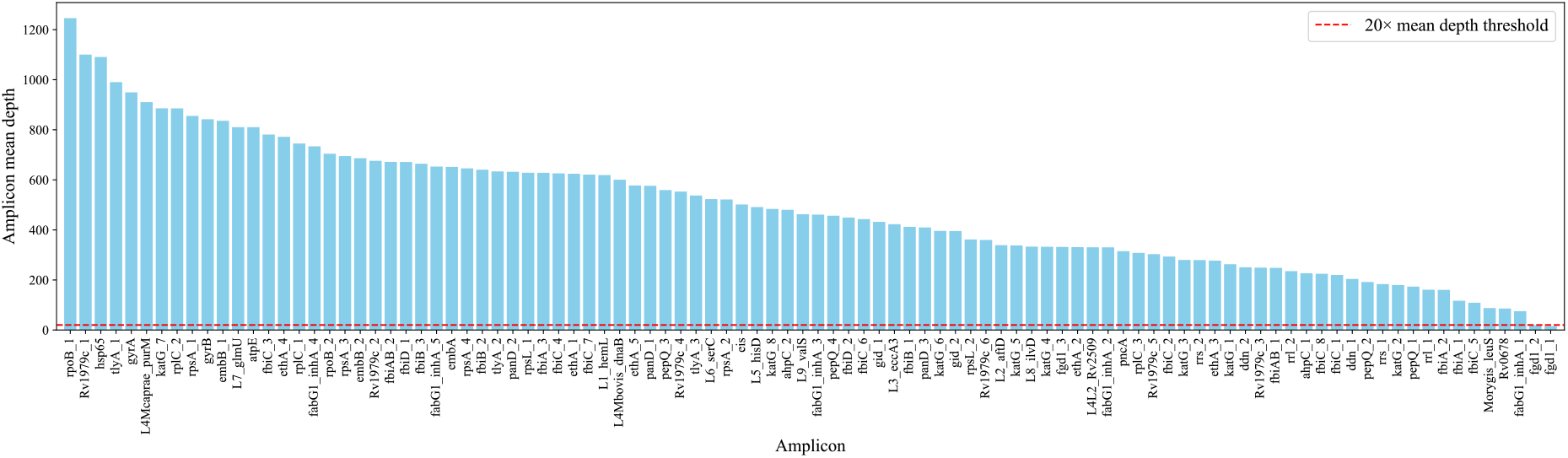
Mean read depth across all 98 tNGS amplicons generated from *M. tuberculosis* H37Rv (DNA concentration: 250 ng/μL). The dashed red line indicates the threshold of 20× mean depth.

**Figure S3:**
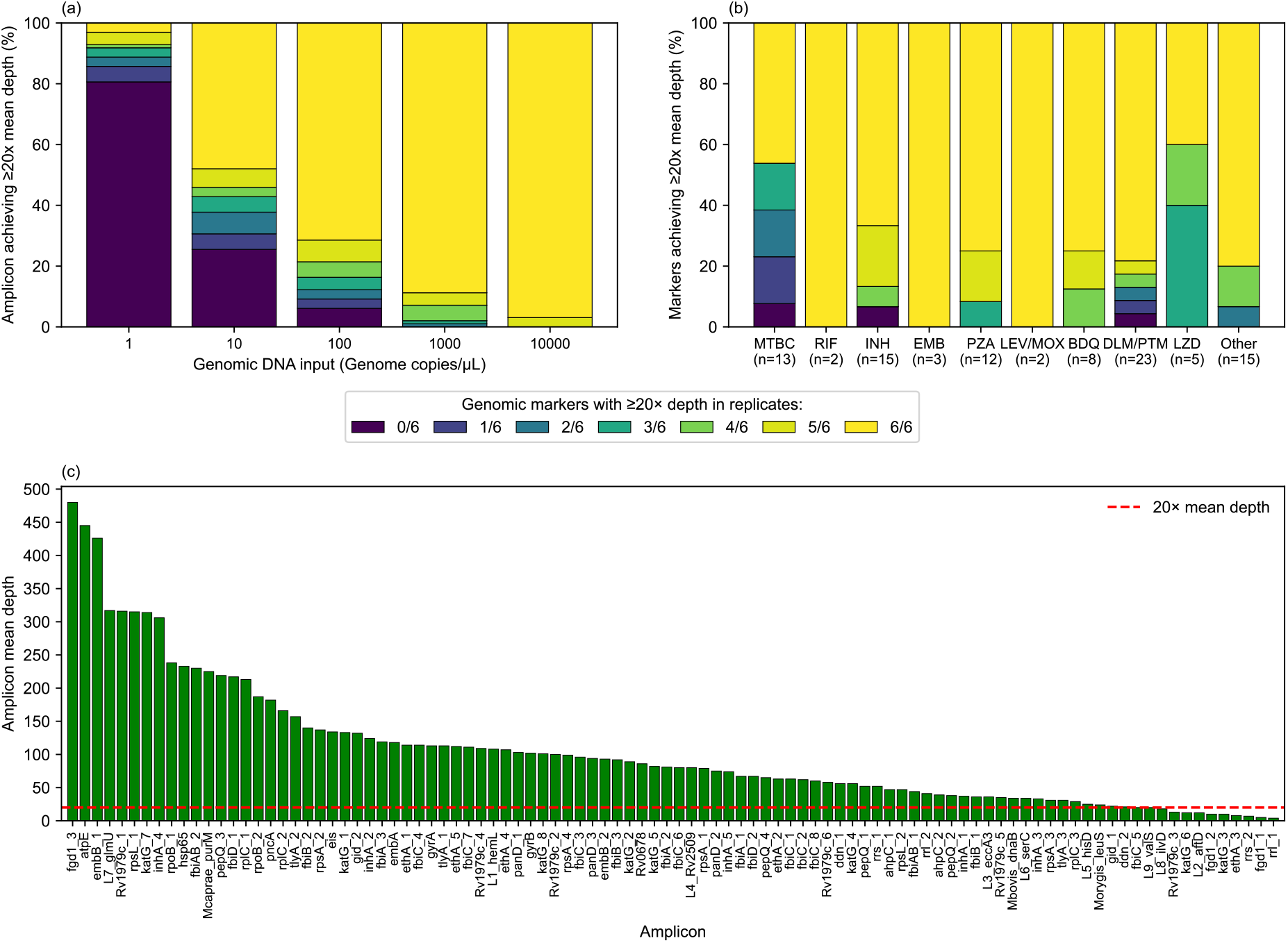
tNGS assay amplicon detection sensitivity with increasing amount of *M. tuberculosis* H37Rv genomic DNA. (a) Detection of 98 amplicons at DNA input levels ranging from 1 to 10,000 genome copies/μL. (b) Detection of 98 amplicons consistently detected at ≥100 genome copies/μL across six replicates. (c) Average of amplicon mean depth at ≥100 genome copies/μL across six replicates. Each bar represents the sequencing mean depth of one of 98 amplicons, aligned to a custom reference of concatenated sequences of 98 designed amplicons. The dashed red line indicates the threshold of 20× mean depth. *Total number of markers evaluated per drug displayed on the x-axis in brackets. BDQ: bedaquiline; DLM: delamanid; EMB: ethambutol; INH: isoniazid; LEV: levofloxacin; LZD: linezolid; MOX: moxifloxacin; MTBC: *Mycobacterium tuberculosis* complex; PTM: pretomanid; PZA: pyrazinamide; RIF: rifampicin; Other: Ethionamide, Amikacin, Capreomycin, Kanamycin, Streptomycin; tNGS: Amplification-based targeted next generation sequencing.

**Figure S4:**
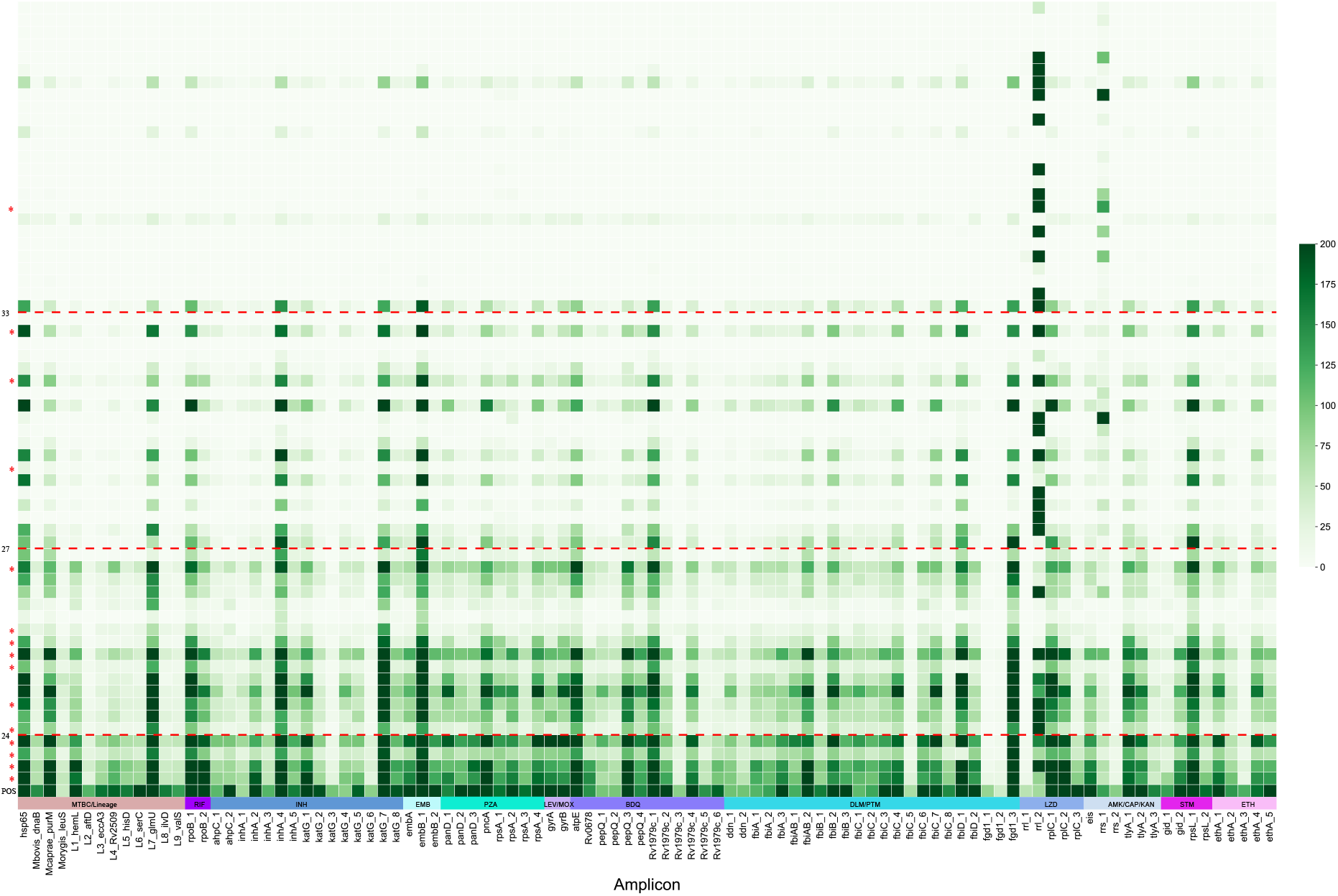
tNGS assay amplicon detection sensitivity in 63 clinical specimens. Each column represents an amplicon, and each row represents a clinical specimen (n=63), ordered by increasing IS*6110* PCR Ct value (decreasing DNA amount). A positive control (POS) of 10,000 genome copies/μL is included - first row. *Smear-positive specimens. Dashed line: IS*6110* PCR Ct value thresholds according to amplicon mean depth ≥20×: • High clinical utility – Ct value ≤24; ≥90% (average 91·8%) • Some clinical utility – Ct value 24·1 - 27: 50-90% (average 63·3%) • Limited clinical utility – Ct value 27·1 - 33: <50% (average 22·8%) • No clinical utility – Ct value >33 AMK: amikacin; BDQ: bedaquiline; CAP: capreomycin; Ct: real-time PCR cycle threshold values targeting IS*6110*; DLM: delamanid; EMB: ethambutol; ETH: ethionamide; INH: isoniazid; IS: Insertion Sequence; KAN: kanamycin; LEV: levofloxacin; LZD: linezolid; MOX: moxifloxacin; MTBC: *Mycobacterium tuberculosis* complex; PCR: Polymerase Chain Reaction; PTM: pretomanid; PZA: pyrazinamide; RIF: rifampicin; STM: streptomycin; tNGS: Amplification-based targeted next generation sequencing.

**Figure S5:**
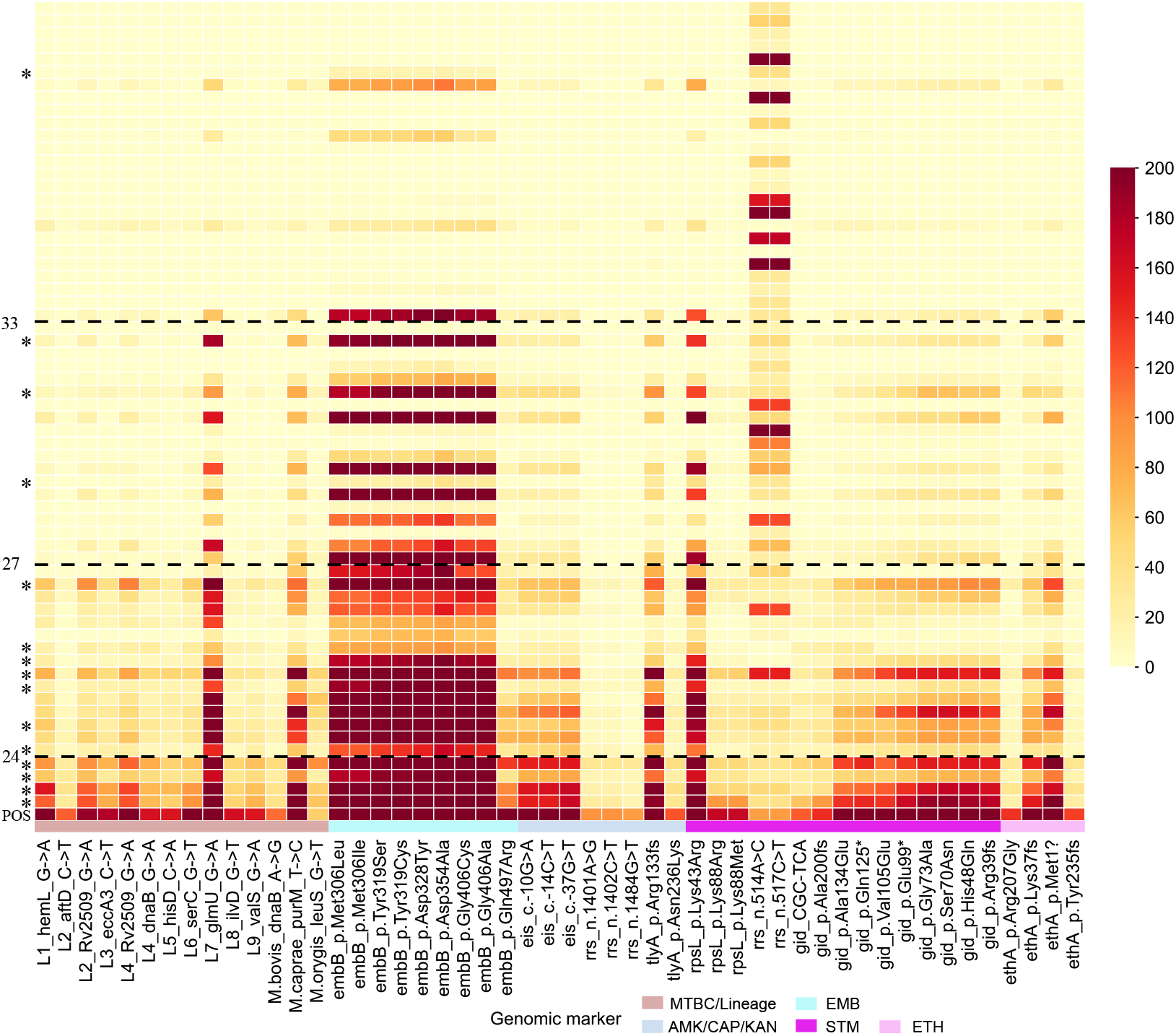
tNGS assay genomic marker detection sensitivity for MTBC/lineages, ethambutol, and other second-line drugs in 63 clinical specimens. Each column represents a drug-resistant marker classified as “Assoc w R” (group 1) in the 2023 WHO catalogue, indicating high clinical relevance. Each row represents a clinical specimen (n=63), ordered by increasing IS*6110* PCR Ct value (decreasing DNA amount). A positive control (POS) of 10,000 genome copies/μL is included - first row. *Smear-positive specimens. Dashed line: IS*6110* PCR Ct value thresholds according to amplicon mean depth ≥20×: • High clinical utility – Ct value ≤24; ≥90% (average 91·8%) • Some clinical utility – Ct value 24·1 - 27: 50-90% (average 63·3%) • Limited clinical utility – Ct value 27·1 - 33: <50% (average 22·8%) • No clinical utility – Ct value >33 AMK: amikacin; CAP: capreomycin; Ct: real-time PCR cycle threshold values targeting IS*6110*; EMB: ethambutol; ETH: ethionamide; IS: Insertion Sequence; KAN: kanamycin; MTBC: *Mycobacterium tuberculosis* complex; PCR: Polymerase Chain Reaction; STM: streptomycin; tNGS: Amplification-based targeted next generation sequencing.

**Figure S6:**
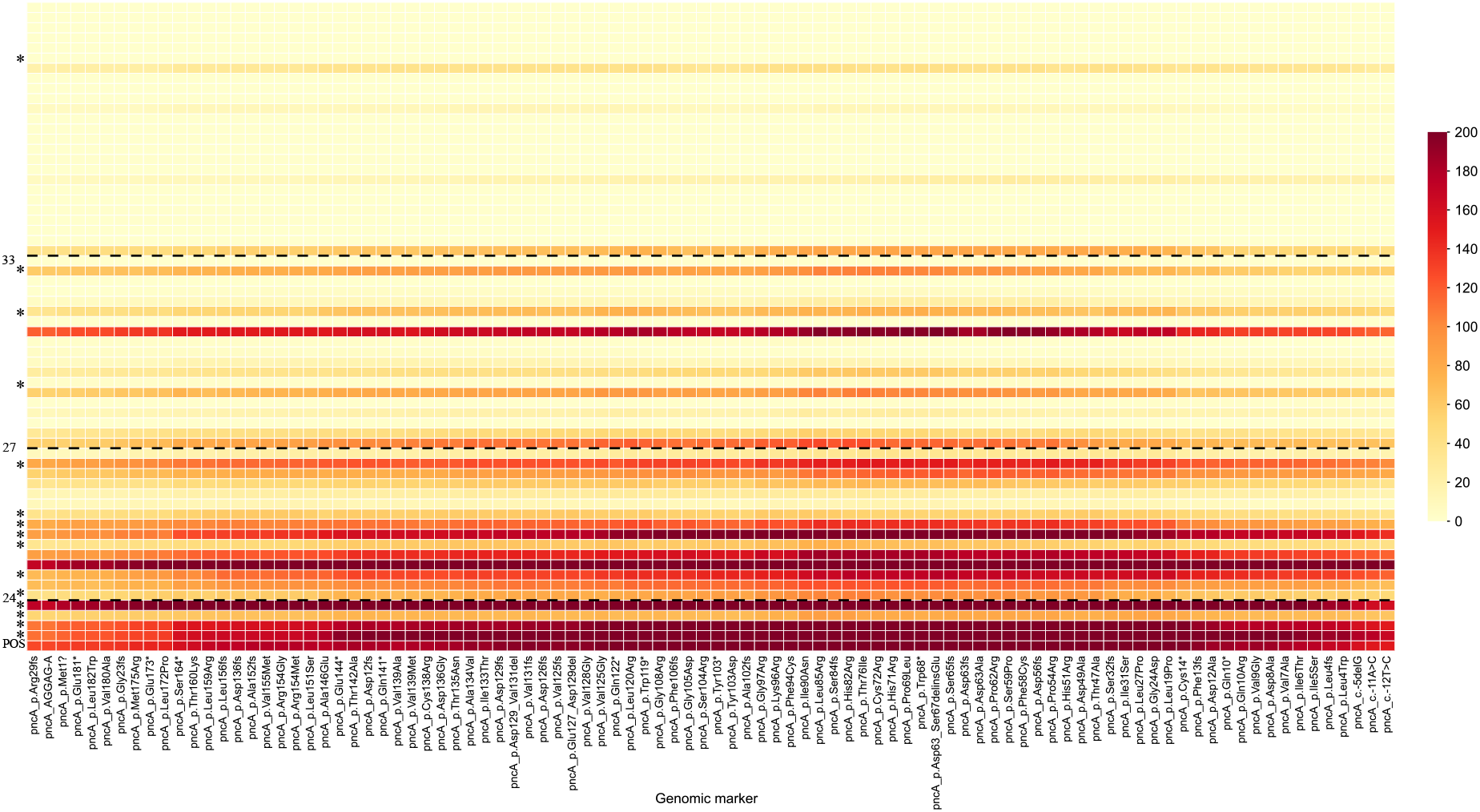
tNGS assay genomic marker detection sensitivity for pyrazinamide in 63 clinical specimens. Each column represents a drug-resistant marker classified as “Assoc w R” (group 1) in the 2023 WHO catalogue, indicating high clinical relevance. Each row represents a clinical specimen (n=63), ordered by increasing IS*6110* PCR Ct value (decreasing DNA amount). A positive control (POS) of 10,000 genome copies/μL is included - first row. *Smear-positive specimens. Dashed line: IS*6110* PCR Ct value thresholds according to amplicon mean depth ≥20×: • High clinical utility – Ct value ≤24; ≥90% (average 91·8%) • Some clinical utility – Ct value 24·1 - 27: 50-90% (average 63·3%) • Limited clinical utility – Ct value 27·1 - 33: <50% (average 22·8%) • No clinical utility – Ct value >33 Ct: real-time PCR cycle threshold values targeting IS*6110*; IS: Insertion Sequence; PCR: Polymerase Chain Reaction.

**Figure S7:**
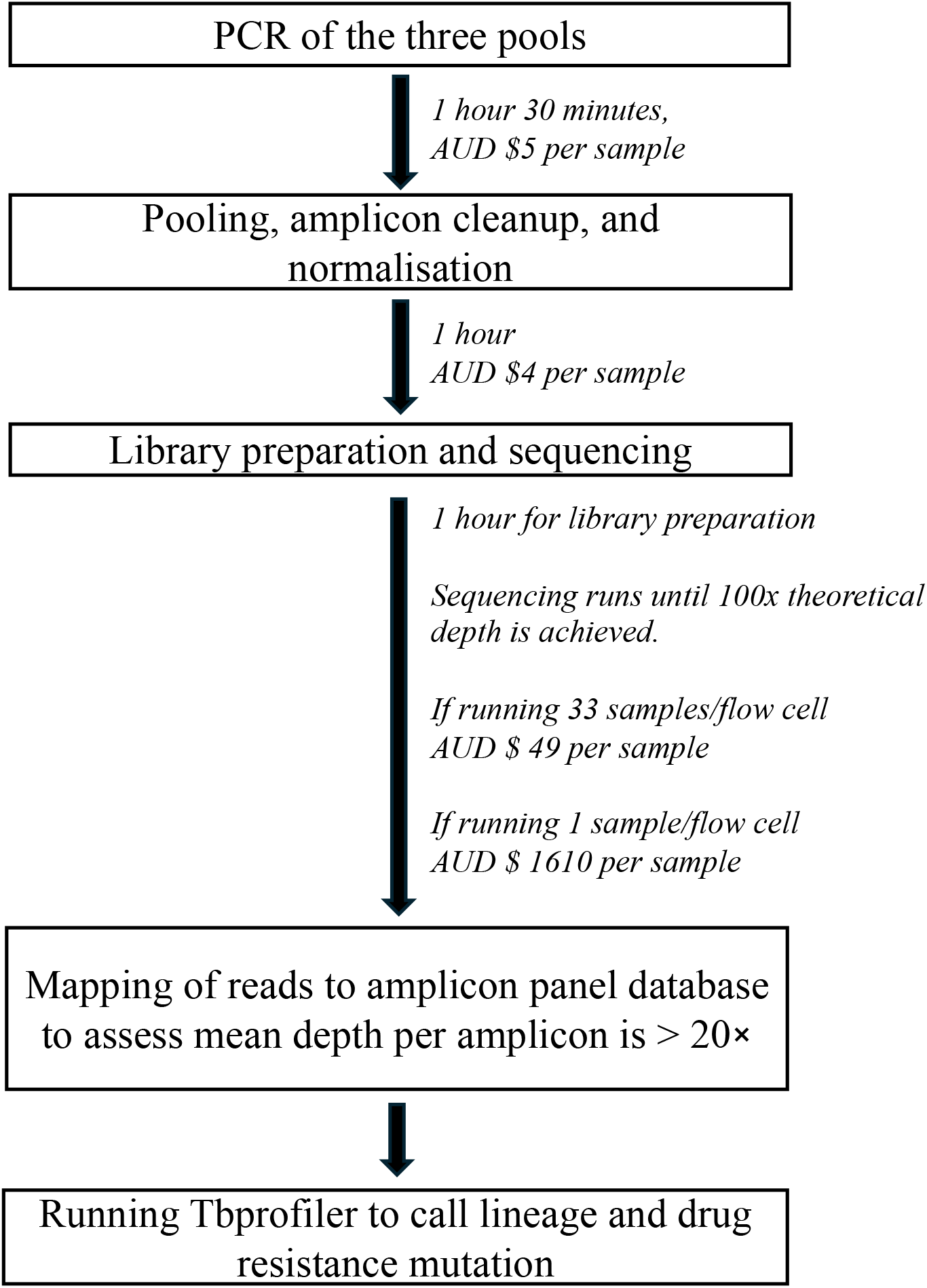
Delineation of the tNGS process. Metrics of either time taken or required read depth are listed for each process. Cost (rounded to the nearest dollar) was calculated from the reagents, kit, and flow cell used in the process. Laboratory plasticware and cost of oligos not included.

